# Novel gain-of-function mutation in dysferlin causes vesicle trafficking defect and IL-1 mediated autoinflammation

**DOI:** 10.64898/2026.08.04.26358821

**Authors:** Farzana Bhuyan, Clinton Bradfield, Amitava Roy, Adriana A. de Jesus, Mohammad Arif Rahman, Benjamin Schwarz, Anjelika Gasilina, Andre Rastegar, Gaurav Sachin, Christopher L. Friend, Kanika Chopra, Kat Uss, Ryan Kissinger, Sara Alehashemi, Sundar Ganesan, Nathan T. Brandes, Ian S. Lacroix, Vinod Nair, Jacqueline M. Leung, Clayton Winkler, Juraj Kabat, Steven M. Holland, Philip J. Kahn, Douglas B. Kuhns, John Hammer, Ronit Herzog, Deborah Consolini, Iain Fraser, Raphaela Goldbach-Mansky

## Abstract

De novo mutations underlying early-onset systemic autoinflammatory diseases have identified key regulators of innate immunity, including pathways that drive IL-1-mmediated inflammation. Here we describe two unrelated girls presenting in infancy with systemic inflammation and sterile lung abscesses, who harbor the same de novo gain-of-function mutation in dysferlin (*DYSF*; p.P1449L) Myeloid expression of *DYSF* P1449L enhances COP-I binding, promotes dysferlin retention in the ER-Golgi, and disrupts vesicle trafficking and membrane homeostasis. Dysferlin-mutant monocytes and M2-like macrophages exhibit ectopic perinuclear NLRP3 inflammasome activation, increased IL-1β production, and inflammatory cell death. Mutant M2-like macrophages further display defects in membrane expansion, exocytosis, efferocytosis, and debris clearance, promoting neutrophil recruitment and DAMP-signal amplification that culminate in sterile abscess formation. These findings identify dysferlin as a regulator of membrane homeostasis in myeloid cells, establish defective membrane-stress adaptation as trigger of NLRP3 inflammasome activation, and define a novel IL-1 mediated autoinflammatory disease caused by gain-of-function *DYSF* mutations.

## Introduction

Sterile inflammation in early childhood often presents as an acute infection-like illness, with fever and systemic inflammation that can rapidly progress to organ damage and death. Among these disorders, monogenic IL-1-mediated autoinflammatory diseases represent a key subgroup defined by excessive IL-1 production and/or signaling^1^ and are highly responsive to IL-1 blockade. Early diagnosis is therefore critical to prevent irreversible tissue injury, yet is often delayed because clinical features, including fever, systemic inflammation and even sterile abscesses, are often indistinguishable from infection. This overlap underscores the need to consider autoinflammation in the differential diagnosis of children with persistent inflammation despite antibiotic therapy and, when appropriate, administer anti-inflammatory treatment.

Genetic studies of early-onset autoinflammation have shown that both de novo gain-of-function mutations and recessive loss-of-function defects in metabolic or mitochondrial stress pathways can drive sterile inflammation, most commonly through activation of the NLRP3 inflammasome^2^. These disorders have provided key insights into how cellular stress pathways converge on IL-1β maturation and release. However, mechanisms linking membrane stress to inflammasome activation remain incompletely defined^3^.

Here we studied two patients who developed antibiotic-refractory sterile lung abscesses and systemic inflammation in infancy following minor respiratory infections, both of whom responded to chronic IL-1 blockade. Both patients carried the same de novo mutation in *DYSF*, isoform 7 (*DYSF-7)* p.P1449L, prompting investigation beyond its established role in muscle. Although biallelic loss-of-function mutations in *DYSF* cause muscular dystrophy through defects in myoblast fusion and membrane repair^4–6^, our patients lacked a muscle phenotype. Instead, our analyses identify dysferlin as a regulator of lipid transport, membrane integrity, and vesicular trafficking in myeloid cells. Disruption of this pathway promotes ectopic NLRP3 inflammasome activation and IL-1β release, defining a non-muscle inflammatory phenotype of dysferlin dysfunction and expanding the spectrum of membrane stress pathways driving IL-1-mediated autoinflammation. More broadly, these findings illustrate how genetic analysis of patients with severe early-onset inflammatory phenotypes can uncover unexpected regulators of innate immune homeostasis.

## Results

### A de novo *DYSF* GOF mutation causes sterile lung abscesses and systemic inflammation in in 2 unrelated infants

We performed trio whole-exome sequencing (WES) in two patients with early-onset sterile inflammation who presented with unexplained fever and, systemic inflammation (elevated C-reactive protein (CRP) and leukocytosis), culture-negative lung abscesses and necrotic gingivitis unresponsive to broad-spectrum antibacterial and antifungal therapy (Fig.1a-c and Extended Data Fig.1a-d). WES identified the same identical de novo germline mutation in exon 39 impacting the C2E domain of dysferlin, a type II transmembrane protein involved in vesicle formation and trafficking^7^, (*DYSF-7*, c.4346C>T, p.P1449L; transcript ENST00000409582.7), (Fig. 1b, d and Extended Data Fig.1a, e, f). In peripheral blood, dysferlin expression was highest in neutrophils, high in monocytes and minimal in lymphoid or dendritic cell populations (Fig. 1e). Both patients developed sterile abscesses at sites of antibiotic injection but showed no evidence of muscle disease (Extended Data Fig. 1b, c and Supplementary note 1). The variant was absent from public databases, including gnomAD (Supplementary table 1). Clinical features suggestive of IL-1-driven inflammation led to empirical treatment with IL-1 blockade, resulting in marked clinical improvement in both patients.

**Fig. 1.**
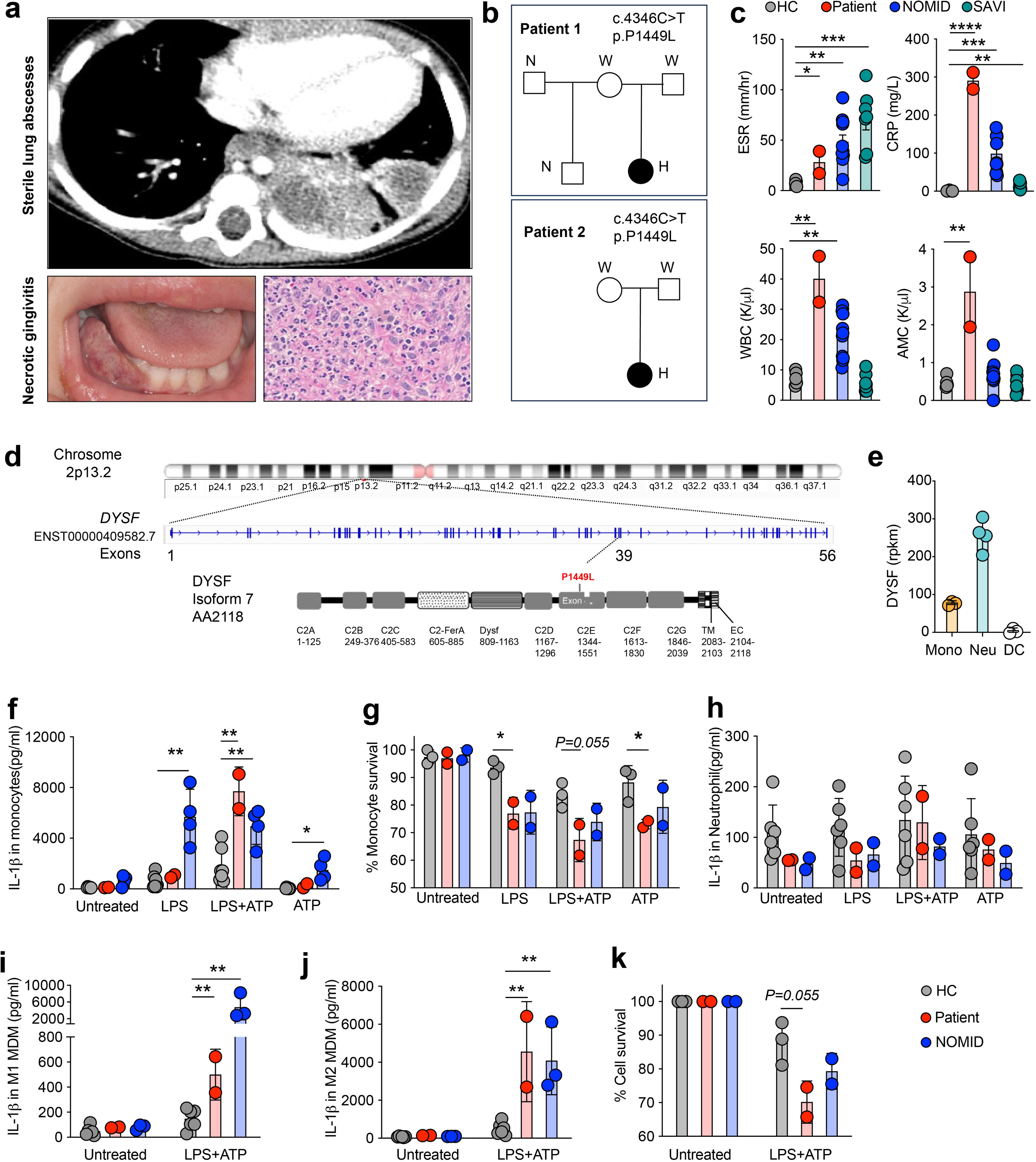
A de novo gain-of-function *DYSF* mutation causes IL-1 mediated systemic inflammation and sterile lung abscesses. (a) Chest radiograph from patient 1 at presentation demonstrating sterile lung abscesses, with representative images of necrotizing gingivitis and gingival biopsy showing neutrophilic infiltration. (b) Pedigrees of two unrelated patients carrying the same de novo *DYSF* mutation. (c) Both patients presented with systemic inflammation and recurrent fever, increased WBC counts, absolute monocyte counts (AMCs), C-reactive protein (CRP), and erythrocyte sedimentation rate (ESR). (d) Domain organization of the blood-expressed dysferlin(*DYSF*) isoform indicating the mutation site. (e) Dysferlin expression in monocytes, neutrophils, and dendritic cells. (f) Patient and control adherent monocytes were stimulated with LPS (1 μg/ml, 2.5 h) followed by ATP (3 mM, 30 min); IL-1β in supernatants was measured by ELISA and compared with healthy controls and patients with NOMID. (g) Cell viability was assessed by MTT assay in ± LPS/ATP-stimulated monocytes after 3 h of stimulation. (h) Neutrophils were stimulated as in f, and IL-1β was quantified by ELISA. (i,j) Monocytes were differentiated for 7 d into M1 or M2-like macrophages, stimulated with LPS and ATP for 24 h, and analyzed for IL-1β secretion. (k) M2-like macrophage viability was assessed by MTT assay before and after 24 h stimulation. Data show mean ± s.d.; dysferlin-mutant monocyte, neutrophil, and macrophage values represent averages from two independent visits with technical replicates. NOMID samples served as disease comparators for inflammasome-associated IL-1β responses in each assay. Significance was determined by unpaired t-test or Mann-Whitney test; *P* < 0.05.

### Dysferlin mutation enhances IL-1β production and cell death in monocytes and MDMs

To assess IL-1β production in myeloid cells, we isolated monocytes and neutrophils from the two dysferlin-mutant patients, healthy controls (HC) and, as disease controls patients with Neonatal-onset multisystem inflammatory disease (NOMID) as inflammatory disease controls. NOMID is caused by gain-of-function *NLRP3* mutations that drive constitutive inflammasome activation^8^. Following co-stimulation with LPS and ATP, dysferlin-mutated patients’ monocytes showed increased levels of cleaved IL-1β, similar to that seen in NOMID, with increased cell death. These are consistent with cell-intrinsic inflammasome activation. Dysferlin-mutant patient-derived monocytes had increased dysmorphic nuclei compared to NOMID (Fig. 1f, g and Extended Data Fig. 2b). Upon LPS and ATP stimulation dysferlin colocalizes with NLRP3 to form puncta. In HC and NOMID monocytes, dysferlin-positive puncta were distributed throughout the cytoplasm and extended toward the cell periphery, whereas in dysferlin-mutant cells dysferlin was markedly reduced in peripheral area (Exended Data Fig. 2a). By contrast, the dysferlin mutation did not increase IL-1β production in patient neutrophils upon LPS and/or ATP stimulation (Fig. 1 h) and did not alter monocyte polarization in blood. However, CD163 expression was increased in classical and intermediate monocytes, indicating persistent activation despite IL-1 blockade and normal inflammatory markers at the time of blood draw (Extended Data Fig. 2c-e).

**Fig. 2.**
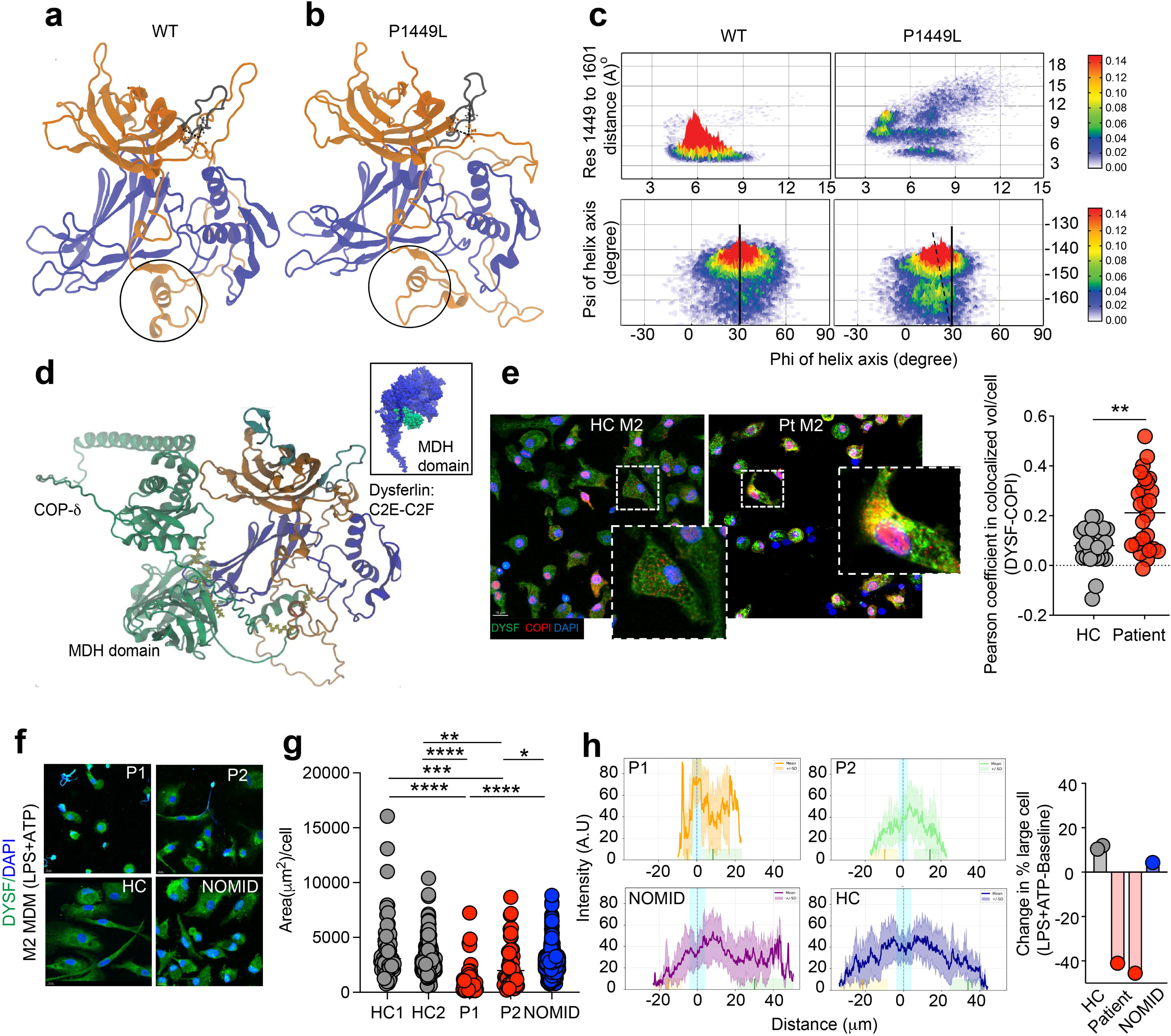
Structural modelling predicts altered dysferlin conformation and COPI interactions. **(a)** Ribbon model of the dysferlin C2E and C2F domains. The green circle highlights the conserved proline residue (P1449), whereas the black circle denotes the α-helix connected to the distal proline-rich region through an unstructured linker. **(b)** Structural modelling of wild-type and mutant proteins. In the wild-type structure, AA P1449 remains in close proximity to two neighboring proline residues, forming a proline stack (blue circle). Substitution of P1449 with leucine (L) weakens this interaction. The wild-type proline (P) occupies the stacked conformation approximately 95% of the time, compared with ∼30% for the histidine variant and ∼14% for the leucine variant. **(c)** Analysis of the helix orientation demonstrates a shift in the backbone φ-φ angle of the α-helix highlighted in **a** (black circle). In the wild-type structure, the helix is maintained at approximately 30°, whereas the leucine variant shifts by ∼15° to approximately 15°, exposing adjacent lysine-rich motifs. **(d)** Structural modelling predicts an interaction between the MHD domain of COPI δ and the C2E-C2F region of dysferlin, with binding localized to a di-lysine-rich region of dysferlin; animated modelling supports stable association of COPI δ with this region. **(e)** Monocytes from healthy controls and dysferlin-mutant patients were differentiated for 7 d into M2-like macrophages, stained for DYSF, COPI and DAPI, and analyzed by immunofluorescence. (f,g) Pearson colocalization coefficients were quantified using Imaris. (h) Single cell profiling of M2 macrophages from healthy-controls, patients and NOMID stimulated with LPS&ATP for 24 h and stained for dysferlin and DAPI. Significance was assessed by unpaired t-test or Mann-Whitney test; *P* < 0.05.

We next generated monocyte-derived M1/M2-like macrophages (MDM) from the dysferlin-mutant patients, HC, and NOMID controls to understand whether dysferlin mutations cause an intrinsic inflammatory defect in macrophages. Dysferlin-mutant macrophages showed increased IL-1β production, most prominently in M2-like macrophages, in which levels were comparable to those in NOMID-derived M2-like macrophages (Fig. 1i, j). LPS- and ATP-induced cell death was also increased in dysferlin-mutant M2-like macrophages relative to control and NOMID M2-like macrophages (Fig. 1k).

### Mutant P1449L dysferlin leads to dysferlin retention, enhanced COP-I/COPδ binding and disruption of membrane dynamics

Dysferlin is a transmembrane protein containing seven canonical C2 domains. To characterize the structural consequences of the mutation, we modeled full-length dysferlin and focused on the C2E-C2F region harboring the P1449L variant. Molecular dynamics simulations revealed that the P1449L substitution disrupts a proline stack, reduces proximity to neighboring prolines and increases flexibility of the adjacent disordered region. This reorients a downstream α-helix (residues 1516-1525), exposing a di-lysine–rich binding interface (Fig. 2a-c and Extended Data Fig. 3a, b). This helix contains a WxxF motif (residues 1516-1519) and a KKxx motif (residues 1518-1520), both implicated in binding the coat protein complex I (COPI), which mediates retrograde Golgi-to-ER transport. While the α- and β-subunits of the COPI complex, recognize the KKXX motifs^9^, the δ-COP subunit binds WxxF motifs^10^. To assess these interactions, we generated AlphaFold models of dysferlin-COPI complexes. In the top five models, δ-COP consistently interacted with the C2E-C2F helix (Fig. 2d, Extended Data Fig.3c and Supplementary Video 1), whereas COPα or COPβ showed no interaction (Extended Data Fig 3d and Supplementary Note 2). Consistent with these predictions, immunostaining of patient-derived M2-like MDMs demonstrated some perinuclear colocalization of dysferlin and COPI (Fig. 2e). These findings indicate that mutant dysferlin increases COPI binding, promoting ER-Golgi retention. Accordingly, patient-derived M2-like MDMs were predominantly small and exhibited restricted distribution, with dysferlin signal concentrated near the nucleus and a steep decline towards the cell periphery (Fig. 2f-g). Following stimulation, the proportion of large cells increased in healthy controls (by ∼15%) and remained relatively stable in NOMID macrophages (increased by ∼10%), whereas it decreased by 40-50% in dysferlin-mutant cells. Together, these findings demonstrate quantitative and qualitative differences in cell size and dysferlin distribution across experimental groups (Fig. 2h).

### Impaired membrane dynamics disrupt efferocytosis, podosome formation, exocytosis, and cell spreading

To determine whether the dysferlin mutation contributes to sterile lung abscess formation through defective neutrophil clearance, we first examined homeostatic neutrophil turnover, focusing on CXCR4-dependent homing of aged neutrophils to the bone marrow^11–13^ (Extended Data Fig. 4a). Although total CXCR4 expression was reduced in patient neutrophils, surface expression and peripheral blood neutrophil counts remained normal (Extended Data Fig. 4b, c), indicating intact homeostatic trafficking.

We next assessed efferocytosis, a key pathway for clearance of apoptotic neutrophils at sites of inflammation^14^, to distinguish neutrophil-from macrophage-intrinsic effects. HC macrophages engulfed both control and patient neutrophils with similar efficiency (Fig. 3a-d and Extended Data Fig. 4d, e), whereas, patient-derived macrophages showed markedly impaired efferocytosis, most prominently in M2-like MDMs (Fig. 3b). In a 17-h assay, efferocytosis reached ∼70% in HC but only ∼30% in patient cells (Fig. 3c and Supplementary Video 2), indicating a macrophage-intrinsic defect, consistent with impaired clearance of apoptotic neutrophils in sterile lung abscess formation^15^.

**Fig. 3:**
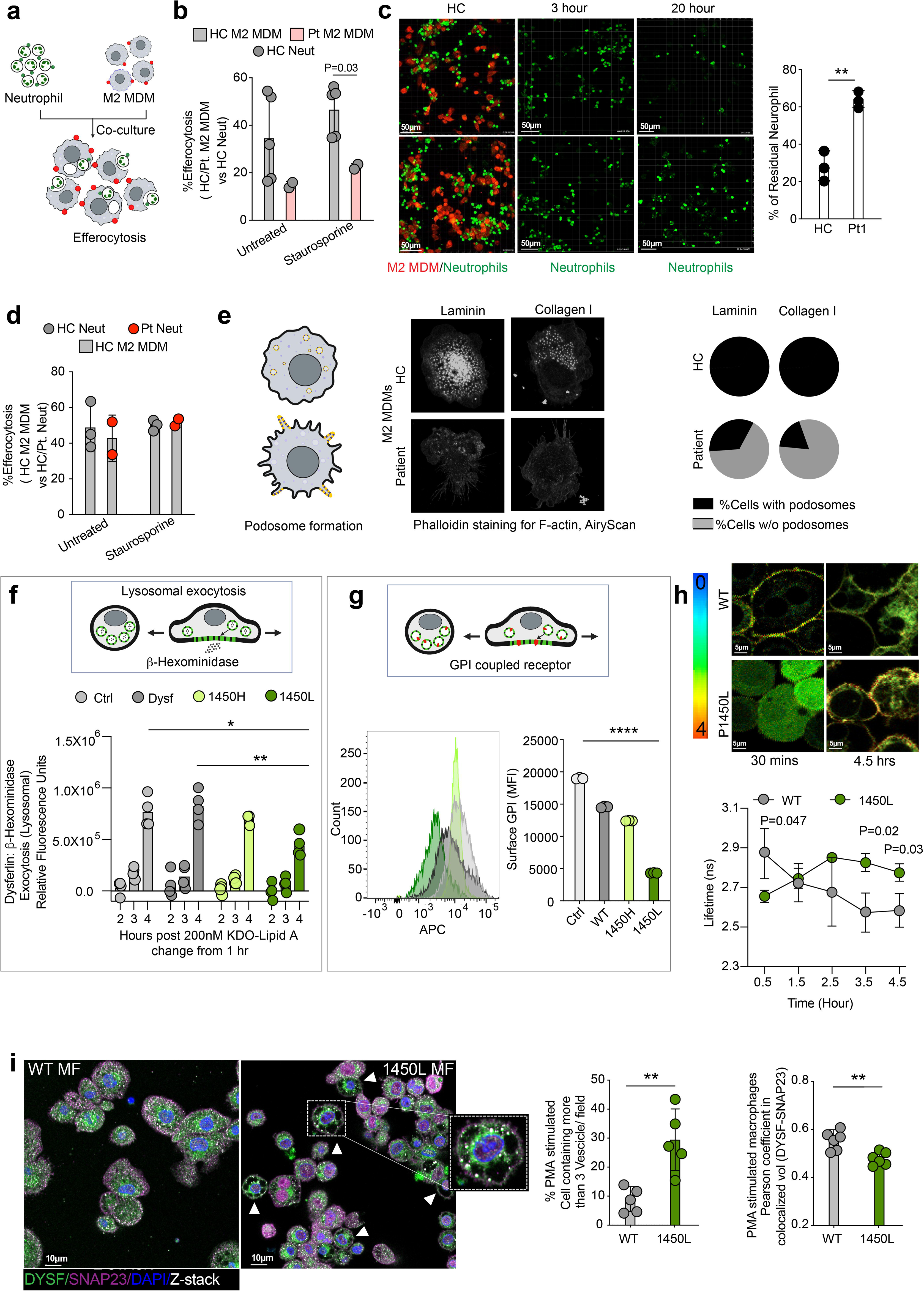
Impaired efferocytosis caused by dysregulated membrane dynamics, spreading, and defective podosome formation. (a) Schematic of the efferocytosis assay. M2-like monocyte-derived macrophages (M2 MDMs) were labeled with Tag-it Violet dye, and neutrophils were labeled with CFSE before co-culture. (b) M2 MDMs differentiated for 7 d from patient or healthy-control monocytes were co-cultured for 3 h with healthy-control neutrophils treated with or without staurosporine. Double-positive cells were quantified by flow cytometry to measure efferocytosis. (c) Patient and control M2 MDMs co-cultured with healthy-control neutrophils were monitored by fluorescence lifetime imaging microscopy (FLIM) for 17 h after the initial 3 h co-culture. (d) Patient neutrophils were labeled with CFSE, treated with or without staurosporine, and co-cultured with healthy-control M2 MDMs; double-positive cells were quantified after 3 h across two independent patient visits. (e) M2 MDMs were plated on glass-bottom chambers coated with laminin, fibronectin or collagen I. F-actin was stained with phalloidin and imaged by Airyscan confocal microscopy; pseudocolor indicates signal intensity. Podosome formation was quantified across the different extracellular substrates. (f) U937 cells expressing wild-type or mutant dysferlin were differentiated with PMA and M-CSF, rested for 24 h, and assessed for lysosomal exocytosis by β-hexosaminidase release and immunofluorescence. (g) Surface GPI expression was measured by flow cytometry. (h) PMA-derived macrophages were stimulated with LPS and ATP, labeled with Flipper probe, and imaged by FLIM. (i) Representative DYSF and SNAP-23 immunofluorescence images are shown. Data represent values from technical replicates where indicated. Significance was determined by unpaired t-test or Mann-Whitney test; *P* < 0.05.

Because spreading and efferocytosis require dynamic membrane remodeling^16^, we next examined membrane organization and trafficking. Podosome formation was markedly reduced in patient-derived macrophages, across all matrices tested (Fig. 3e and Extended Data Fig. 4f). In U937 cells stably expressing the disease-associated P1450L dysferlin variant (annotation in isoform 13 (*DYSF-13)* corresponnding to P1449L in *DYSF-7*), but not wild-type dysferlin or the non-pathogenic P1450H variant, we observed impaired spreading, reduced podosome formation increased COP-1 colocalization and decreased bead phagocytosis (Extended Data Fig. 5a-f).

As membrane expansion during cell spreading depends on exocytosis^17,18,19,20^ we assessed vesicular trafficking and membrane tension. Lysosomal and GPI-associated exocytosis were significantly reduced in P1450L*-*expressing cells despite preserved lysosomal acidification (Fig. 3f, g and Extended Data Fig. 5f). Flipper-TR^21^ measurements showed delayed (∼2.5 h) and sustained (4.5h) membrane tension in mutant cells (Fig. 3h) and impaired transcritpional upregulation of *DYSF* in stimulated M2-MDMs compared to NOMID (Supplementary Note 3), indicating impaired resolution of membrane tension and impaired membrane delivery.

Because dysferlin also promotes SNARE-mediated vesicle docking and fusion through interaction with SNAP23 and syntaxin-4^22–25^, we next examined membrane fusion. Dysferlin staining was also increased in perinuclear membrane compartments, consistent with retained membrane intermediates (Extended Data Fig. 6a, b) and dysferlin-lined enlarged vesicle-like structures accumulated under the plasma membrane and showed reduced dysferlin-SNAP23 colocalization (Fig. 3i) consistent with impaired SNARE-mediated membrane fusion. In parallel, plasma membrane phosphatidylserine (PS), required for efficient SNARE-mediated membrane fusion, was reduced in both mutant U937 cells and patient-derived M2-like macrophages (Extended Data Fig. 6c-e). Together, these findings indicate that the dysferlin mutation disrupts membrane expansion, vesicle trafficking, and exocytosis, resulting in defective efferocytosis and broad defects in membrane homeostasis.

### Mutant dysferlin links lipid dysregulation to maladaptive stress responses and compartmentalized inflammasome activation in M2-like MDMs

Given impaired membrane fusion and other defects in cell structure, we next examined lipid homeostasis. To determine whether the dysferlin mutation alters lipid homeostasis in hematopoietic cells, we compared the lipidomes of PBMCs from HC (biological n = 3, technical n=6) and patients (biological n = 2, technical n=6) following LPS+ATP stimulation. Lipidomic profiling revealed dysregulation of multiple lipid classes, including phosphoglycerols (PGs), lysophospholipids (LPC), and glucosylceramides (GlcCers) (Fig. 4a-c and Extended Data Fig. 7a-c). PG species showed altered composition consistent with disrupted ER-mitochondria lipid handling (Fig. 4a) whereas LPC levels were reduced (Fig. 4b), consistent with impaired vesicular lipid delivery and reduced membrane curvature. In parallel, glycosphingolipids were decreased (Fig. 4c), indicating impaired sphingolipid processing and transport in the Golgi apparatus and/or membrane raft assembly^26^. Altered acyl-chain composition, characterized by accumulation of intermediate species (e.g., C22:4) and reduced C22:5 (DHA), further suggested disrupted ER-peroxisome lipid flux and impaired peroxisomal processing. (Fig. 4d and Extended Data Fig. 7d).

**Fig. 4:**
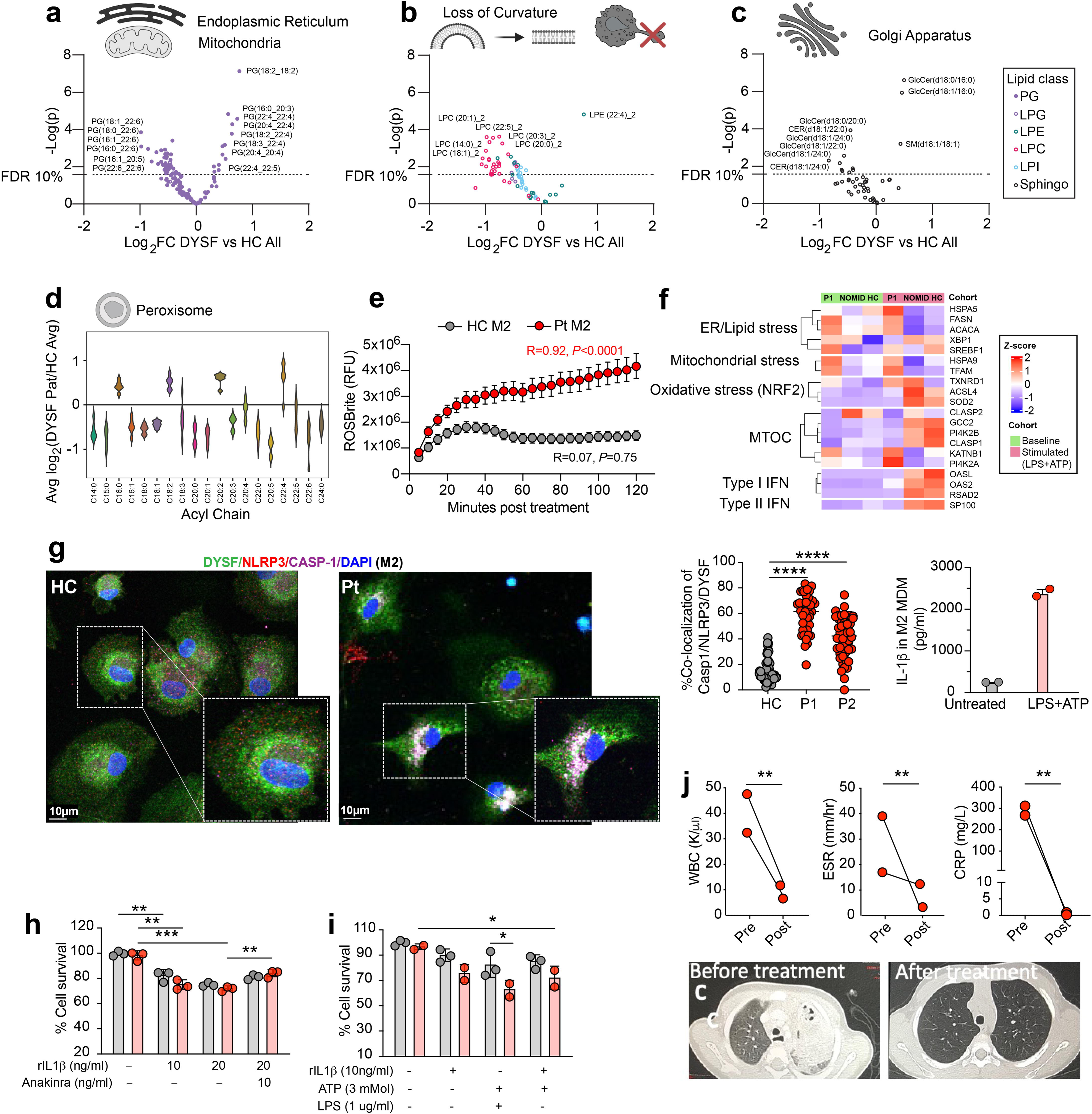
Mutant dysferlin disrupts of lipid homeostasis, increases membrane tension, and promotes ectopic inflammasome activation. **(a-c)** Lipidomic profiling of resting PBMCs from a DYSF-mutant patient and healthy controls. Selected lipid species are shown from the comprehensive analysis in Extended Data Fig. 7c; technical replicates (n = 6) were analyzed for the patient and each of three healthy controls. **(a)** Endoplasmic reticulum- and mitochondria-associated phosphatidylglycerol (PG) species differentially abundant in DYSF patient PBMCs compared with healthy controls. **(b)** Curvature-inducing and stress-associated lysophospholipid species identified in the same analysis as in **(a). (c)** Differentially abundant sphingolipids, including Golgi-derived glycosphingolipids, identified in the same analysis as in (a). **(d)** Average log base-2-fold change of lipid abundance in patient PBMCs relative to healthy controls, grouped by the most unsaturated acyl chain within each lipid species to assess global acyl-chain remodeling. **(e)** Total reactive oxygen species production in patient-derived and healthy-control M2-like monocyte-derived macrophages after LPS and ATP stimulation. **(f)** Bulk RNA-sequencing of M2 macrophages from patient P1(n=1), NOMID controls(n=3) and healthy controls(n=2) at baseline and after LPS/ATP stimulation, showing pathway activity related to ER/lipid stress, mitochondrial stress, cytoskeletal and MTOC organization, and type I and II interferon signaling as log2 fold change relative to healthy controls. **(g)** Representative immunofluorescence images of patient-derived M2 macrophages stained for DYSF(green), NLRP3(red) and caspase-1(purple) after stimulation. (h,i) Effects of IL-1 and anakinra on patient M2 macrophage viability and metabolic activity measured by MTT assay. (j) ESR, CRP and WBC counts before and after anakinra, with radiographic resolution of lung abscesses following therapy. significance was determined by Unpaired-t-test or Mann-Whitney test. * p <0.05

Because altered lipid metabolism can impact redox homeostasis, we next assessed ROS production. Following LPS+ATP stimulation, total ROS was markedly increased in dysferlin-mutant M2-MDMs but only minimally in M1 MDMs (Fig. 4e and Extended Data Fig. 7e), indicating lineage-specific susceptibility to oxidative stress. Transcriptomic analysis revealed that dysferlin-mutant M2-like MDMs adopt a maladaptive stress state characterized by enhanced ER/lipid stress and mitochondrial stress responses, accompanied by impaired activation of adaptive programs regulating calcium homeostasis, autophagy, oxidative stress responses, mitochondiral maintenance, and cytoskeletal organization. These abnormalities became more pronounced following LPS+ATP stimulation and included dysregulation of genes involved in microtubule organization, vesicular trafficking, and nuclear-cytoskeletal architecture, including *CLASP1/2, KATNB1, PI4K2A/B, GCC2*, and *SYNE2*. In contrast to NOMID macrophages, which largely preserved coordinated stress-adaptation pathways, DYSF-mutant M2 MDMs exhibited impaired induction of stress-response networks involved in autophagy and calcium homeostasis (*MTMR3, ORAI1, ITPR3*), oxidative and metabolic adaptation (*NFE2L1, HIF1A*), and mitochondrial maintenance. Collectively, these findings indicated a failure to mount coordinated organelle stress-adaptation programs downstream of defective lipid trafficking (Fig. 4f and Extended Data Fig. 7f).

To define the subcelluar origin of IL-1β production, we examined NLRP3 inflammasome activation in dysferlin-mutant macrophages. Under basal conditions, NLRP3 was diffusely distributed throughout the cytoplasm with minimal colocalization with caspase-1 (Fig. 4g). Following LPS+ATP stimulation, NLRP3 and caspase-1 redistributed to perinuclear compartments in dysferlin-mutant M2-like MDMs but not in M1 MDMs, resulting in increased colocalization compared to HC and NOMID controls (Fig. 4g). This phenotype was not observed in M1 MDMs and was absent following NLRC4 staining of M2-MDMs (Extended Data Fig. 8a, b). NLRP3, caspase-1, and dysferlin partially overlapped within these perinuclear regions and were associated with increased IL-1β production (Fig. 4g). In parallel, dysferlin-mutant M2-like MDMs exhibited increased numbers of dysmorphic nuclei, extracellular NLRP3-positive as well as NLRP3/dysferlin-positive puncta (Extended Data Fig 8c-d), consistent with release of inflammasome-associated membrane material during inflammatory cell death. Dysferlin-mutant M2-like MDMs also selectively induced CCL2 and CXCL1 following stimulation (Extended Data Fig. 9a, b), a response not observed in NOMID M2-MDMs despite constitutive NLRP3 activation. By contrast, stimulated dysferlin-mutant neutrophils released higher levels of IL-16 and MIF (Extended Data Fig 9c), cytokines associated with inflammatory cell death and secondary necrosis. Recombinant IL-1β induced dose-dependent cell death in dysferlin-mutant macrophages, which was partially rescued by IL-1 blockade (Fig. 4h), indicating that IL-1 signaling amplifies macrophage dysfunction. Consistent with these findings, IL-1 blockade normalized inflammatory markers, resolved sterile lung abscesses, and prevented recurrent inflammatory flares in both patients (Fig 4i), whereas interruption of the IL-1 inhibition was associated with recurrence of severe necrotizing lung inflammation requiring intensive care support (Supplementary Case description P2). Together, these findings indicate that dysferlin-mutant M2 macrophages exhibit maladaptive stress responses associated with ectopic perinuclear inflammasome activation, selective chemokine induction and amplified IL-1 signaling, supporting a central role for IL-1 in sustaining this inflammatory state (Extended Data Fig 9d).

## Discussion

A single heterozygous de novo missense mutation in dysferlin (p.P1449) defines a previously unrecognized monogenic autoinflammatory disease characterized by IL-1-mediated systemic inflammation, defective efferocytosis, and sterile lung abscess formation. Dysferlin is a member of the ferlin family, an evolutionarily conserved group of membrane-fusion proteins implicated in membrane repair, vesicle trafficking, and phospholipid homeostasis,^27,28^ however, the role of ferlins in immune-cell membrane homeostasis has remained incompletely understood. Through disease-based discovery, we identify a previously unappreciated role for dysferlin in myeloid cells as a regulator of membrane trafficking and membrane stress adaptation linked to ectopic inflammasome activation. In contrast to canonical inflammasomopathies such as NOMID, in which inflammasome activation results directly from gain-of-function mutations in *NLRP3*, the dysferlin p.P1449 mutation appears to confer a gain-of-function interaction with COP-I/COPδ that disrupts vesicle trafficking and lipid transport required for membrane homeostasis. Rather than directly activating the inflammasome, dysferlin dysfunction creates a maladaptive membrane stress state characterized by retention of dysferlin-positive membrane compartments that serve as sites of ectopic inflammasome assembly. While NLRP3 activation may initially represent an appropriate stress response, the resulting IL-1-driven program generates a self-amplifying inflammatory loop, that in the presence of defective neutrophil clearance, perpetuates rather than resolves tissue injury.

Dysferlin, a type II transmembrane protein, containing multiple calcium-sensitive C2 domains,^29^ has classically been linked to sarcolemmal membrane repair in muscular dystrophies^25^ and also interacts with proteins involved in SNARE-dependent membrane fusion and vesicle trafficking^23,30^. Dysferlin also binds phosphatidylserine (PS) and other lipids in a Ca²-dependent manner^30^ and has been implicated in regulating cellular lipid composition^31^, including PS levels through endosomal and lysosomal fusion^32^. Our findings extend these functions to myeloid cells and suggest that dysferlin-dependent membrane remodeling is particularly critical in M2-like tissue macrophages during inflammatory resolution. Professional phagocytes require continuous membrane remodeling to sustain inflammatory responses and efferocytic clearance during infection and tissue injury^33,34^, when large numbers of neutrophils, apoptotic cells, and inflammatory debris must be cleared to restore tissue homeostasis. The preferential involvement of M2-like macrophages likely reflects their role as the principal efferocytic phagocytes responsible for neutrophil clearance and tissue repair. Although our data implicate defective macrophage efferocytosis as a major driver of disease, activated neutrophils can also contribute to clearance of dying neutrophils during inflammation^15,35^, and whether this compensatory pathway is impaired in dysferlin-mutant neutrophils will be an important topic for future investigation.

Our data support a model in which dysferlin-mutant macrophages fail to appropriately adapt to this increased efferocytic burden. Retention of dysferlin-positive membrane compartments near ER/Golgi-associated regions likely reflects impaired trafficking and defective mobilization of membrane reservoirs required for phagocytic expansion and inflammatory adaptation. Under homeostatic conditions these defects may remain partially compensated; however, during inflammatory stress, impaired membrane remodeling limits the ability of macrophages to clear accumulating neutrophils and dying cells. The resulting unresolved membrane and organelle stress promotes ectopic perinuclear inflammasome assembly (Fig 4g), oxidative and ER stress, and mitochondrial dysfunction (Extended Data Fig 7f), ultimately shifting macrophages from a reparative toward a pathologic inflammatory state.

NLRP3 inflammasome assembly has previously been linked to phosphatidylinositol-4-phosphate (PI4P)-enriched membranes of the trans-Golgi network following stimulation^36,37^ and our findings are consistent with a model in which dysregulated membrane organization creates aberrant membrane platforms permissive for inflammasome assembly. Dysferlin, NLRP3, and caspase-1 partially colocalized within perinuclear membrane compartments, supporting a close spatial relationship between retained vesicular intermediates and inflammasome activation. Importantly, dysferlin-mutant macrophages differed substantially from NOMID macrophages despite comparable IL-1β production. Whereas NOMID macrophages retained relatively preserved adaptive stress responses, dysferlin-mutant macrophages exhibited persistent membrane stress, oxidative stress, chemokine induction, and defective inflammatory adaptation, suggesting that the inflammatory phenotype is driven not simply by excess inflammasome activation, but by failure to restore membrane homeostasis during inflammatory stress.

Additional observations warrant further investigation. We identified extracellular NLRP3-positive puncta, some colocalizing with dysferlin and others lacking dysferlin, released from activated and dying macrophages. Because extracellular inflammasome particles can propagate inflammasome activation following uptake by bystander myeloid cells^38,39^, dysferlin-coated puncta may represent a distinct mechanism for sequestration and clearance of inflammasome-associated material compared with uncontained extracellular NLRP3 debris. In parallel, dysferlin-dense puncta accumulated beneath the plasma membrane in activated NOMID M1 and M2 macrophages but were markedly reduced in dysferlin-mutant cells, raising the possibility that dysferlin-containing vesicles contribute to membrane repair during inflammatory pore formation. Failure of this adaptive membrane response may increase susceptibility to pyroptotic cell death in dysferlin-mutant macrophages compared with NOMID macrophages, despite comparable IL-1 production.

Collectively, our study supports a model in which defective membrane adaptation during increased phagocytic and efferocytic demand impairs neutrophil clearance and promotes unresolved membrane stress, ectopic NLRP3 inflammasome activation, and release of inflammasome-associated DAMPs, thereby establishing a self-amplifying IL-1-driven inflammatory cycle. Recombinant IL-1β directly induced macrophage cell death, which was partially rescued by IL-1 blockade, whereas interruption of IL-1 inhibition in patients resulted in recurrence of severe necrotizing lung inflammation that rapidly resolved upon treatment reinstatement, illustrating the central role of IL-1 blockade in maintaining inflammatory homeostasis despite persistence of the underlying membrane-trafficking defect. Our findings identifes dysferlin as a critical regulator of membrane stress adaptation during efferocytosis, revealing an essential role for dysferlin-mediated membrane trafficking in phagocyte adaptation to inflammatory stress and expanding the spectrum of IL-1-mediated autoinflammatory diseases to include disorders of membrane homeostasis in myeloid cells.

## Methods

### Study participants

All research investigations were conducted under protocol 17-I-0016/NCT02974595, that was approved by the NIAID and NIAMS/NIDDK Institutional Review Boards. Patients were referred to the NIH between 2013 and 2015. Written informed consent was obtained from the parents of all patients/healthy controls involved, and assent was obtained from the patients/healthy controls were indicated. Both patients were referred for unexplained systemic inflammation and sterile lung abscesses and underwent trio analysis at the NIH Clinical Center. Clinical, laboratory and imaging data and unstained slides from clinically indicated skin, gastrointestinal and liver biopsies were obtained and stained for research markers. The data collection period for some parameters is from birth to April 2022 for both patients. The NIH protocol enrolls parents and healthy siblings as controls. The control experiments were conducted with blood from the patients’ parents and or controls from the NIH Blood Bank approved under the protocol. The authors affirm that the patients (or their parents/legal guardians) provided written informed consent for publication of the images in the Fig 1 and Extended Data Fig. 1 and the medical information included in this paper. Anakinra was initially started as clinical treatment. Canakinumab was later initiated once patients were in inflammatory remission and had normalized their inflammatory parameters on an off-label basis. The study was reported according to CARE guidelines and conducted in compliance with the Declaration of Helsinki principles.

### Genetic and functional analyses

The primary objectives of the natural history study were to identify the genetic cause in patients with early-onset severe systemic inflammation, and to characterize the disease pathogenesis. We use trio WES and WGS and targeted genetics to identify and validate a genetic discovery and cellular models to mimic and interrogate immune dysregulation induced by a novel mutation in a clinically relevant tissue or cell model. Genomic studies are detailed in the Supplementary Information file.

### Monocyte Isolation, Differentiation of primary human Macrophages (M1/M2 MDMs)

Human peripheral blood derived neutrophils, monocyte and monocyte-derived macrophages (MDMs) were prepared as described previously. Heparinized venous blood was collected from healthy donors after informed consent. For Monocyte isolation and Macrophage culture, PBMCs were suspended in RPMI 1640 medium containing a low concentration of FCS (1%) to facilitate the adherence of monocytes in the subsequent step and then seeded in multiwell plates or dishes. Monocytes were enriched by allowing attachment to plates, chamber slides, or dishes for 1 h at 37°C, and nonadherent cells were removed by extensive wash with PBS. The adherent monocytes were differentiated into macrophages by culturing with RPMI 1640/10% FCS containing 10 ng/ml recombinant human (rh) GM-CSF (Peprotech USA) for M1 MDMs and 100 ng/ml recombinant human (rh) M-CSF (Peprotech USA) for M2 MDMs as previously described ^40^. After 3 d, the cultures were replaced with fresh complete media after extensive wash with PBS to further remove nonadherent cells and were incubated for another 3 d. At day 6, the purity of MDMs prepared by this method was routinely >95% according to flow cytometric analysis of CD14 expression (data not shown).

### Generation of Dysferlin mutant U937 cell line

Production of U937 cells stably expressing dysferlin mutants was performed by making MIG-series retroviral constructs. Briefly, virus was produced by transfecting 293T17 cells with a 3:1:0.1 ratio of EcoPak (Addgene), construct, and VSV-G (Addgene) using Mirus Lenti (). After 72 h, virus was collected and 0.45 um filtered and was directly applied to log-phase U937 cells. After 72 hours, cells were sorted by GFP expression using a FACS Aria and the upper quartile of expressors were use for subsequent experiments. U937 cells were cultured in RPMI containing 10% heat-inactivated FBS and 2mM glutamine and were differentiated in 20ng/mL PMA for 2 days for macrophages. Related to supplementary Data 1

### Cytokine analysis (cytokine array and ELISA)

The relative levels of multiple cytokines and chemokines in the supernatants of macrophages were analyzed using a human cytokine array (R&D Systems) according to the manufacturer’s instructions. In brief, culture supernatants (100 μl), which were collected after centrifugation, were added to dot blots onto which the capture Abs had been spotted in duplicate. After incubation with the secondary Ab mixture, the resultant signals were detected using the Biorad image analyzer. The intensity of the spots was quantified using the Image J software. The supernatants of Neutrophils, Monocytes and M*acrophages* stimulated with or without LPS and ATP were collected and analyzed for the concentration of IL-1b by ELISA (R&D System DY201).

### Immunofluorescence /Image acquisition and analysis

Stimulated and unstimulated monocyte, macrophages and U937 cells transduced to express mutant Dysferlin were fixed in 4% paraformaldehyde in coverslip or in ibidi chamber slides, permeabilized with 0.1% Triton X-100, and stained with a single antibody (Ab) or combinations of anti-NLRP3(Polyclonal; LS-B4321) from LS Bio (Lynwood, WA, USA), Anti-Caspase-1 (Polyclonal; PA5-87536) from Fisher Scientific (Hampton, NH, USA),, anti-Snap23 Ab (EPR8538; Cat. # ab-131242), anti-COP1 Ab (1E4; Cat. # ab-56400) from Abcam (Cambridge, MA, USA), Fluor® 647-anti-DYSF Antibody (Polyclonal; Cat. # bs-2429R-BF647) from Bioss (Woburn, Massachusetts, USA) and Anti-Phosphatidylserine Antibody (1H6; 05-719) from Millipore Sigma, (Rockville, MD USA) for 12 hrs followed by AlexaFluor568-anti-mouse IgG (Cat. # A-11004) from Fisher Scientific (Hampton, NH, USA), AlexaFluor488 anti-rabbit IgG (Cat. # Ab 150077) from Abcam (Cambridge, MA, USA).DAPI (Molecular Probes, Waltham, MA, USA) was used to visualize nuclei. Signals were visualized with confocal laser-scanning microscope (Leica SP8, Buffalo Grove, IL, USA). Image processing was performed using the Imaris 10.0.0.app software.

### Single-cell profiling of M2 Macrophages and analysis

For each cell, a complete linear fluorescence profile was generated across the longest cell diameter using Fiji, extending from one plasma membrane boundary to the opposite membrane boundary and passing through the center of the nucleus. This approach enabled assessment of dysferlin intensity distribution across individual cells and comparison among experimental groups. All profiles are aligned at the nucleus center (position 0) and displayed across a consistent X-axis range of -75.21 to +75.21 µm.Before averaging, each cell profile is automatically evaluated and potentially mirrored so that the shortest and brightest path from nucleus center to membrane always appears on the same side (typically the right side of the plot). This orientation procedure ensured that profiles from individual cells were aligned in a biologically meaningful manner rather than according to arbitrary image orientation. Cells were further stratified by size into small and large cell classes using an approximate threshold of 37.8 μm. Individual cell profiles stacked together to show variability across cells. Mean fluorescence intensity profiles were plotted with standard deviation bands to illustrate the average signal distribution and overall variance within each group. In addition, Gaussian-smoothed mean profiles were displayed with standard error of the mean bands to emphasize group-level trends and confidence in the estimated mean profile.

### Efferocytosis assay

M2 MDM were differentiated from monocyte using 100ng/ml M-CSF for 7 days. Cells were washed in PBS and resuspended at 1x 10^6^ cells/mL then 1uL of 5mM solution of Tag-it Violet™ (Cat: 425101; Biolegend) dye/ml of PBS were added and incubated for 20 minutes at 37°C. Cells were washed with cell culture medium containing 10% FBS for 3 times. Neutrophils were isolated from whole blood and resuspended at 5 x 10^6^ cells/mL PBS, 1ul of 5mM CFSE (Cat: C34554; Thermofisher) solution were added and incubated for 20 minutes at 37°C,Cells were then treated with ± Staurosporine(Cat: 81590; Cayman Chemicals). Macrophage and neutrophil were co-cultured or mixed in 1:3 ratio. Incubated in 37C for 3 hr and at the end of the coculture, cells were washed with PBS, fixed with 1% paraformaldehyde in PBS and acquired on a FACSymphony A5 and examined using FACSDiva software (BD Biosciences) by acquiring all stained cells. Data were further analyzed using FlowJo v10.6 (TreeStar, Inc., Ashland, Oregon, USA). The frequency of efferocytotic cells was determined as the frequency of double-positive cells for Tag-it Violet™and CFSE on the Tag-it Violet™ positive Macrophages. Subsequently Co-culture was filmed under microscopy(FLIM) for 17 hours.

### Whole blood monocyte and neutrophil characterization

The frequency and cytokine levels were measured in patient and control whole blood. Cells were phenotyped and were stimulated in 10% RPMI in the presence/or absence of LPS and ATP for 3 hours. Subsequently, Whole blood was stained with Live/Dead Aqua Dye (cat. #L34966, 0.5 μl) from Thermo Fisher, followed by surface staining with the following: Alexa 700 anti-CD3 (SP34-2; cat. #557917, 5μl), Alexa 700 anti-CD20 (2H7; cat. #560631, 5μl), PE-Cy5 anti-CD86 (IT2.2; cat. #555666, 5μl), BUV805 anti-CD14(M5E2; cat. # 612902, 5μl), BV750 anti-CD206 (19.2; cat. # 746891, 5μl), BV650 anti-CD8(RPA-T8; cat. # 563821, 5μl), BUV563 anti-CD163(GHI/61; cat. # 741402, 5μl), BUV661 anti HLA-DR (G46-6; cat. # 612980, 5μl), Alexa 700 anti-CD11b (ICRF44; cat. #557918, 5μl), APC-Cy7 anti-CD16 (3G8; cat. #557758, 5μl), PE-CF594 anti-CD56 (B159; cat. #562289, 5μl), BV786 anti-CD45 (HI30; cat. # 563716, 5 μl) from BD Biosciences (San Jose, California, USA), FITC anti-CD66abce (TET2; cat. # 130-116-522, 5 μl) from Milteny (Gaithersburg, Maryland) and BV510 anti CD15 (W6D3; cat. # 323028, 5 μl) from Biolegend (San Diego, California). Following surface staining samples were treated with Foxp3 / Transcription Factor Staining Buffer from Thermo Fisher to get rid of red blood cells. Samples were acquired on a BD FACSymphony A5 cytometer and analyzed with FlowJo software 10.6. Neutrophils were gated as singlets, live cells, CD45^+^ cells, CD3^-^, CD20^-^, CD8^-^, CD15^+^, CD16^+^, CD14^-^, and CD66abce^+^ cells. Classical monocytes were gated as singlets, live cells, CD45^+^ cells, CD3^-^, CD20^-^, CD8^-^, CD15^-^, HLA-DR^+^, CD14^+^CD16^-^cells. Intermediate monocytes were gated as singlets, live cells, CD45^+^ cells, CD3^-^, CD20^-^, CD8^-^, CD15^-^, HLA-DR^+^, CD14^+^CD16^+^cells. Non-classical monocytes were gated as singlets, live cells, CD45^+^ cells, CD3^-^, CD20^-^, CD8^-^, CD15^-^, HLA-DR^+^, CD14^-^CD16^+^cells.

### Survival assay

Monocyte, Macrophage numbers were monitored using the MTT reagent. Monocytes and Macrophages were cultured on 24-well tissue culture plates under different conditions. MTT (Invitrogen) was added to each well at a final concentration of 0.5 mg/ml, and 0.01 N HCl-isopropanol was added after the incubation for 4 h at 37°C. The absorbance of the wells was measured at 595 nm using microplate reader (Bio-Rad).

### Exocytosis assays and GPI staining

Lysosome exocytosis from U937 cells expressing dysferlin mutants was quantitated by assessing release of b-hexosaminidase according to manufacturer protocols (MET-5095). Briefly, U937 were plated in 384-well format and were differentiated with 20 ng/mL PMA for 2 d. Medium was exchanged to Optimem containing 200 nM KDO-lipid A, the active signaling component of LPS. Once the endpoint was reached, supernatant was removed from the wells and was mixed with b-hexominosidase fluorogenic substrate for 15 m prior to reading ex:365nm em:450nm on a BMG-VantaSTAR plate reader. 2x10^6^ U937 cells stably expressing indicated Dysferlin variants were differentiated for 2d with 20ng/mL PMA in 6-well plates. Cells were lifted and fixed in 4% PFA for 10 minutes. Cells were washed and quenched with TBS. Cells were extracellularly stained with 1:200 mouse anti-GPI (Thermo ma5-34724) for 1 hour in TBS. Cells were secondarily stained with 1:1000 donkey anti-mouse Alexa 647. Cells were washed 3x in PBS and were analyzed in flow using a BD Fortessa on the APC channel. Quantitation of staining was performed with FlowJo.

### Lysosome and Cathepsin B Analysis

2x10^6^ U937 cells stably expressing indicated Dysferlin variants were differentiated for 2d with 20ng/mL PMA in 6-well plates. Cells were stained for desired components via Immunochemistry Technologies (Kit #938) protocol. For lysosome quantitation, cells were lifted and were stained with 1µM Acridine Orange for 30 minutes. For Cathepsin B staining, cells were treated with 10µM Magic Red for 30 minutes. Cells were washed 3x in PBS and were analyzed in flow using a BD Fortessa on the APC channel. Quantitation of staining was performed with FlowJo.

### Podosome formation

Unstimulated or stimulated cells were fixed in pre-warmed 4% PFA in cytoskeletal buffer (10 mM PIPES/25 mM HEPES pH 6.9, 100 mM NaCl, 1 mM EGTA, 300 mM sucrose, 3 mM MgCl2) and stained with phalloidin AlexaFluor 488 (Cell signaling 8878). Imaging of individual cells were acquired on Zeiss 880 AiryScan equipped with a 60X, 1.4 NA objective using system optimized z-stack sectioning. Resulting images were analyzed in Imaris (Bitplane) and Fiji/ImageJ.

### FLIPPER Assay

Membrane tension was measured using Flipper-TR kit (CY-SC020) from Cytoskeleton, Inc.Co, USA. U937 cell and U937 derived macrophages were fixed in 4% paraformaldehyde in ibidi chamber slides. Flipper-TR was diluted in DMSO to make 1mM stock solution. Final concentration used in cell culture medium was 1µM. Cells were incubated at 37°C for 30 minute before imaging. Cells were imaged with standard FLIM microscopes using a 488 nm pulsed laser for excitation and collecting photons through a 600/50 nm bandpass filter.

### ROS assay

Patient M1 and M2 macrophages were plated in 384-well format and were labeled with 10µM DCFDA and 1µg/mL Hoechst 33342 for 10 m. Medium was replaced with complete RPMI lacking phenol red containing 200 nM KDO-Lipid A. Cells were loaded into a CX7 high-content imager equipped with an environmental control chamber. Cells were imaged every 5 m for 3 h. Cell-average intensiometric data were calculated by HCS Studio software following Hoechst nuclear segmentation and expansion of the nuclear ROI to encompass the cell body.

### Lipidomics

#### Sample Preparation

For all LCMS methods LCMS grade solvents were used. All samples were immersed in 0.4 mL of ice-cold methanol during collection. To each sample 0.4 mL of water and 0.4 mL of chloroform were added. Samples were agitated for 30 minutes with refrigeration and centrifuged at 16K xg for 20 min. 400 µL of the bottom (organic) was collected and dried down under vacuum. The sample was resuspended in an equivalent volume of 5 µg/mL butylated hydroxytoluene in 6:1 isopropanol:methanol for targeted bulk lipidomics and bile acid analysis.

#### Liquid Chromatography Mass Spectrometry

Bulk lipidomics was performed as previously described^41^ with a shortened gradient, polarity switching, and scheduled acquisition. A LD40 X3 UHPLC (Shimadzu Co.) and a 7500 QTrap mass spectrometer (AB Sciex Pte. Ltd.) were used for separation and detection. Lipids were separated by class on a Water XBridge Amide column (3.5 μm, 3 mm X 100 mm) with a 9-minute gradient from 100% 5 mM ammonium acetate, 5% water in acetonitrile apparent pH 8.4 to 95% 5 mM ammonium acetate, 50% water in acetonitrile apparent pH 8.0. All lipids were detected using MRMs that leveraged lipid class-conserved fatty acid product ions, headgroup product ions, or neutral loss ions.

All signals were integrated using SciexOS 3.1 (AB Sciex Pte. Ltd.). Signals with greater than 50% missing values were removed and remaining missing values were replaced with the lowest registered signal value. Dataset was total sum normalized and signals with a QC coefficient of variance greater than 30 % were discarded. A Benjamini-Hochberg method for correction for multiple comparisons was imposed where indicated.

### Protein modeling

We generated five structural models of dysferlin isoform 7 (UNIPROT ID O75923-7) from its sequence using the AlphaFold software^42^ installed on the Biowulf cluster at the National Institutes of Health. We took the top-scored conformation along with estimates of prediction reliability (pLDDT) and predicted aligned error (PAE), as described elsewhere^43^. Briefly, pLDDT indicates the quality of prediction for individual residues in a protein domain, while PAE is governed by the prediction of the interaction between domains, consequently indicating the reliability of the relative position of domains in a multidomain protein.

### Bulk RNA-Sequencing

Patient and control derived M1 and M2 macrophages were stimulated with LPS and ATP for 24 hr. Total RNA was extracted, quality assessed and used for bulk RNA-sequencing library preparation.Libraries were sequenced on an Illumina platform, and reads were aligned to the human reference genome. Gene level counts were generated and analyzed for differential expression.

### Statistical analysis

Statistical analyses were performed using GraphPad Prism version10 and RStudio (R4.4.2) unless otherwise specified. Data are presented as mean ± s.e.m. unless indicated otherwise. Comparisons between two groups were performed using two-tailed unpaired Student’s t tests or Mann-Whitney U tests, depending on data distribution. Comparisons among multiple groups were analyzed using one-way or two-way ANOVA with appropriate multiple-comparison correction. Correlation analyses were performed using Pearson or Spearman correlation coefficients as appropriate. Differential gene expression analyses for RNA sequencing data were performed using DESeq2 with false discovery rate (FDR) correction for multiple testing. A two-sided P value <0.05 was considered statistically significant. Details regarding sample size, statistical tests, and significance thresholds are provided in the corresponding figure legends.

## Data Availability

The data supporting the findings of this study are available from the corresponding author upon reasonable request, subject to National Institutes of Health policies and applicable participant privacy and consent restrictions.

## Data availability

All data supporting the findings of this study are provided as Source data with this paper. Further information will be available from the corresponding author on reasonable request.

## Acknowledgements

We thank the patients and healthy donors for their participation in this study and Valerie Mohammed for assistance with patient scheduling at the NIH.

## Author contributions

F.B. and R.G.-M. conceived the study, wrote the manuscript and finalized the figures; F.B. and C.B designed and performed the transfection model, exocytosis assays and cell-line assays with I.F.; A. Roy. performed dysferlin modeling, calculated and analyzed the MD trajectories; A.A.d.-J. coordinated and performed the genomic analyses; F.B. and M.A.R designed the flow cytometry panel, performed efferocytosis assays and characterized whole blood samples; B.S.,N.T.B. and I.S.L performed lipidomic analyses by mass spectrometry; A.G. and J.H. performed podosome formation assays; A.R., G.S., C.L.F. with F.B. processed samples and prepared cells for immunofluorescence experiments; S.A., S.H., K.U, K.C. assisted with data collection and patient scheduling; R.K. prepared graphical presentations of the data; R.H., D.C. and P.J.K. treated the patients and provided clinical follow-up. D.K. assisted with neutrophil characterization; F.B. with C.W. performed the high resolution immunostaining. S.G., J.L., J.K., V.N. assited with data acquisition and analysis. R.G.-M. followed the patient and oversaw the project execution. All authors reviewed and edited the final version of the manuscript.

## Competing interests

The authors declare no competing interests.

## Funding source

This research was supported in part by the Intramural Research Program of the National Institutes of Health (NIH). The contributions of the NIH author(s) are considered Works of the United States Government. The findings and conclusions presented in this paper are those of the author(s) and do not necessarily reflect the views of the NIH or the U.S. Department of Health and Human Services.

## Extended Data Figures

**Extended Data Fig 1.**
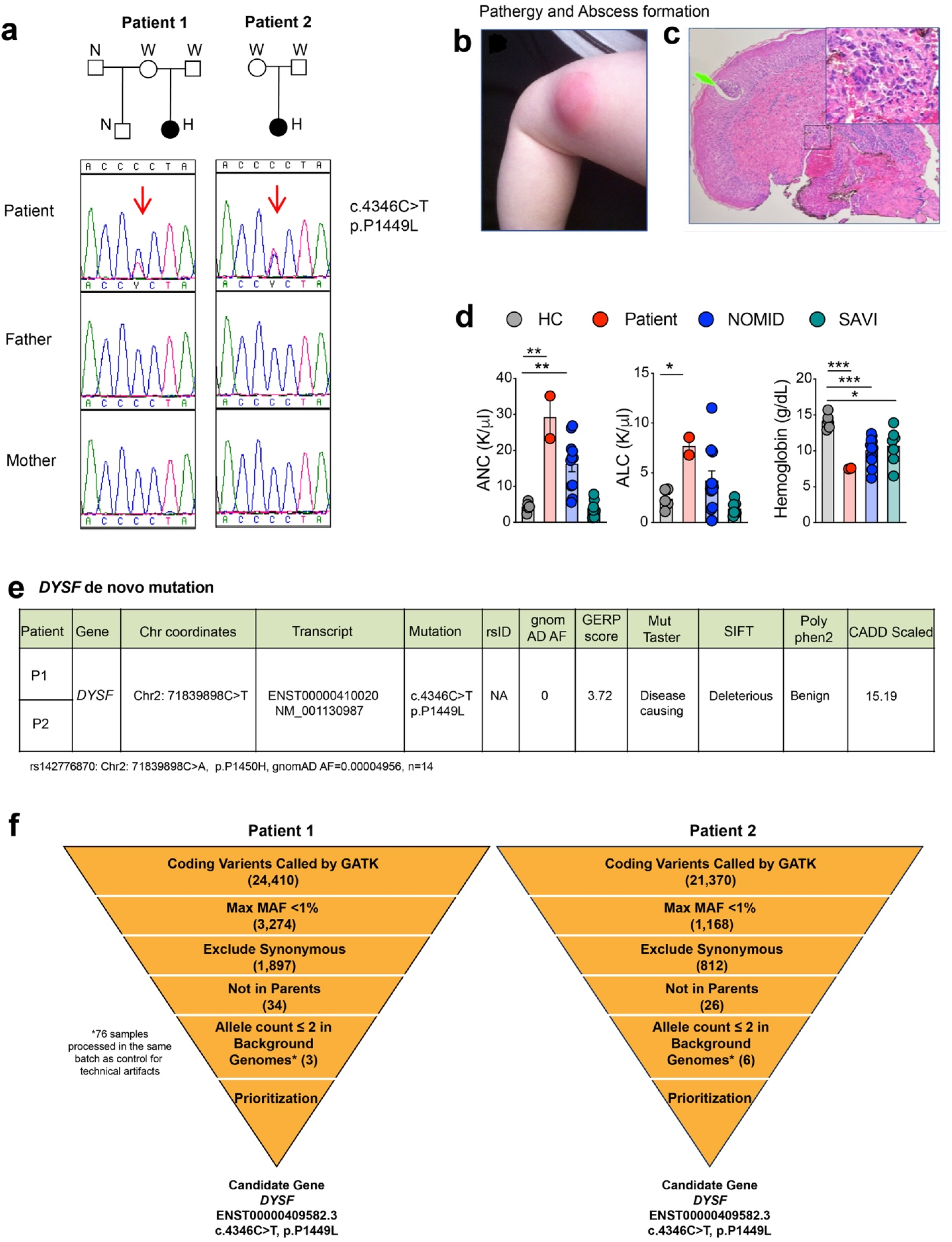
Clinical features and genetic identification of a recurrent de novo *DYSF* mutation. **a,** Both patients carried the same heterozygous de novo mutation in *DYSF*. **b, c,** Representative images demonstrating post injection pathergy with abscess formation and neutrophil infiltration confirmed by skin biopsy of the thigh abscess. **d,** Both patients had elevated absolute neutrophil counts (ANCs) and absolute lymphocyte counts (ALCs), with reduced hemoglobin levels. **e,f,** Whole exome sequencing identified the same de novo germline mutation in exon 39 of *DYSF* c.4346C>T, p.P1449L; transcript ENST00000409582.7 affecting the C2E domain of dysferlin.

**Extended Data Fig 2.**
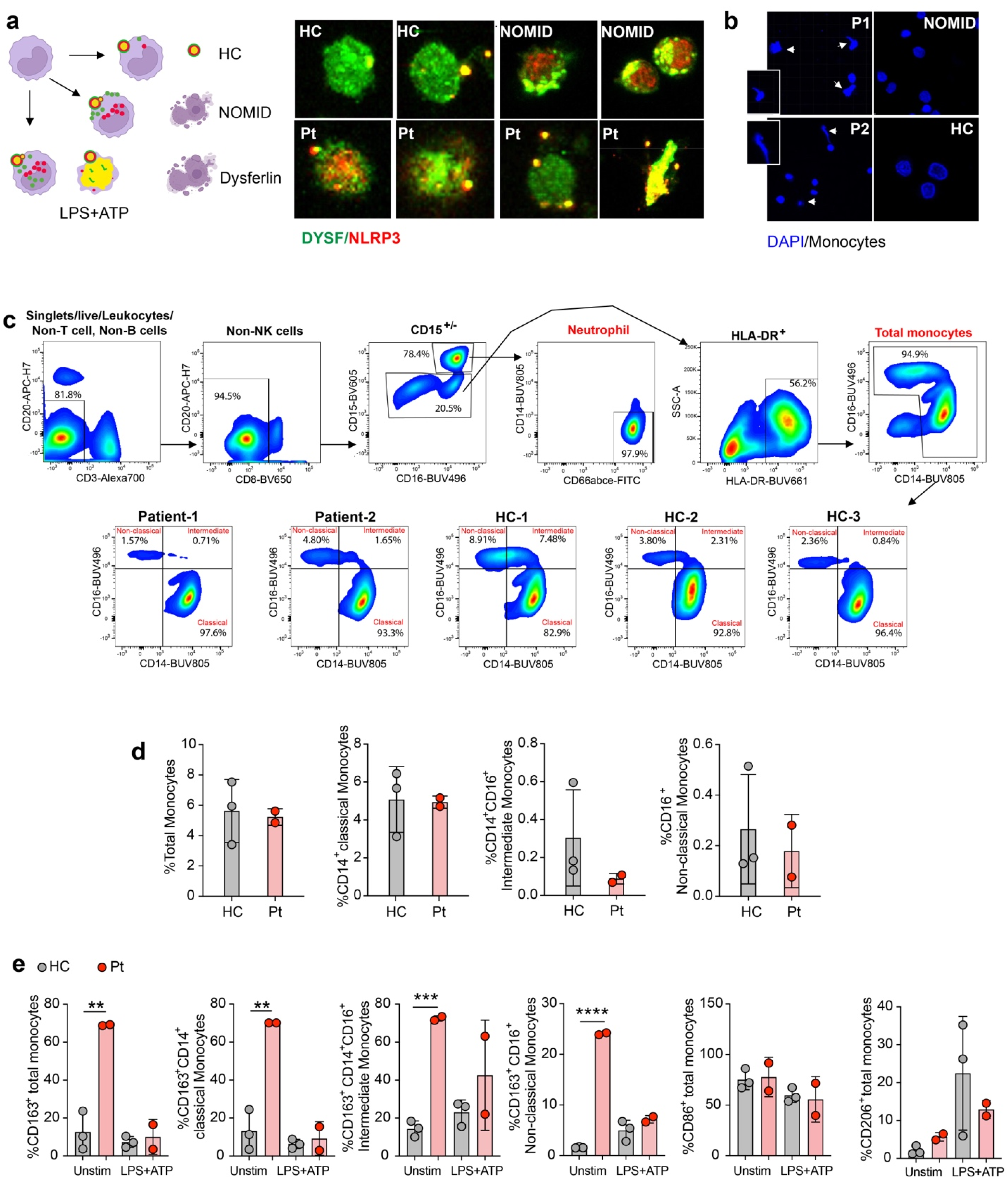
Dysferlin-mutant monocytes and macrophages exhibit nuclear dysmorphism, enhanced NLRP3 puncta formation and increased CD163 expression. **a**, **b,** Adherent PBMC-derived monocytes were stimulated with LPS (1 μg ml/ml, 2.5 h) followed by ATP (3 mM, 30 min), then stained for NLRP3 (red), dysferlin/DYSF (green) and DAPI. Representative cropped immunofluorescence images from patient, healthy control and NOMID monocytes demonstrate distinct patterns of NLRP3 puncta formation with increased numbers of NLRP3 puncta in NOMID and dysferlin-mutant monocytes, accompanied by increased nuclear dysmorphism in dysferlin-mutant patients. **c,d,** Whole blood monocyte frequencies and phenotypes were assessed after culture in RPMI containing 10% serum, with or without LPS/ATP stimulation for 3 h. **e,** The dysferlin mutation did not alter the proportions of classical, intermediate or non-classical monocytes, but CD163 expression was increased across monocyte subsets, most prominently in classical and intermediate monocytes.

**Extended Data Fig 3.**
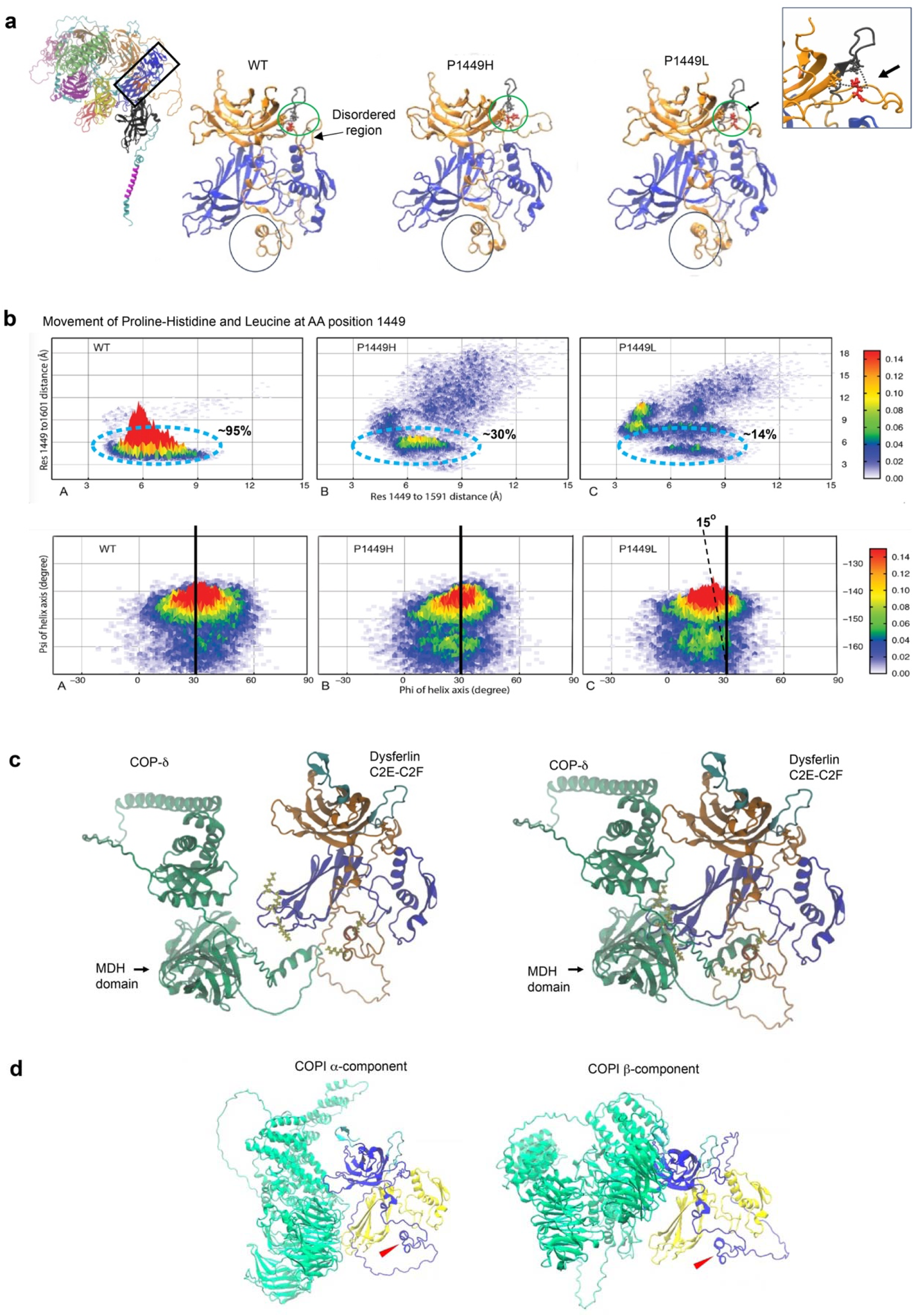
Structural modelling of dysferlin and predicted COPI interactions. **a**, Ribbon model of the dysferlin C2E and C2F domains. Shown for comparison is a rare population variant, p.P1449H (dbSNP rs142776870; ClinVar 2036638), identified in gnomAD (allele frequency, 0.0001171). The green circle highlights the proline stack centered on residue P1449, whereas the black circle denotes an α-helix connected to the distal proline-rich region through a disordered linker. **b**, Molecular dynamics (MD) simulations revealed that interactions among residues P1449, P1591 and P1601 stabilize the conformation of a disordered region spanning residues 1449-1516. In the wild-type protein, P1449 remains in close proximity to P1591 and P1601 approximately 95% of the time. In contrast, the p.P1449H variant maintains this interaction only ∼30% of the time, whereas the disease-associated p.P1449L variant does so only ∼14% of the time. Loss of the proline stack increases conformational flexibility and alters the orientation of the α-helix formed by residues 1516-1525, most prominently in the p.P1449L variant, resulting in exposure of a di-lysine-rich interface predicted to mediate COPI binding. **c,** Structural modelling predicts an interaction between the COPI δ MHD domain and the dysferlin C2E-C2F region through the exposed di-lysine-containing binding interface**. d,** Dynamic modelling of the dysferlin C2E-C2F domains predicts binding of COPI α and COPI β subunits at structurally conserved regions of dysferlin that are spatially distinct from the site affected by the disease-causing p.P1449L mutation.

**Extended Data Fig 4.**
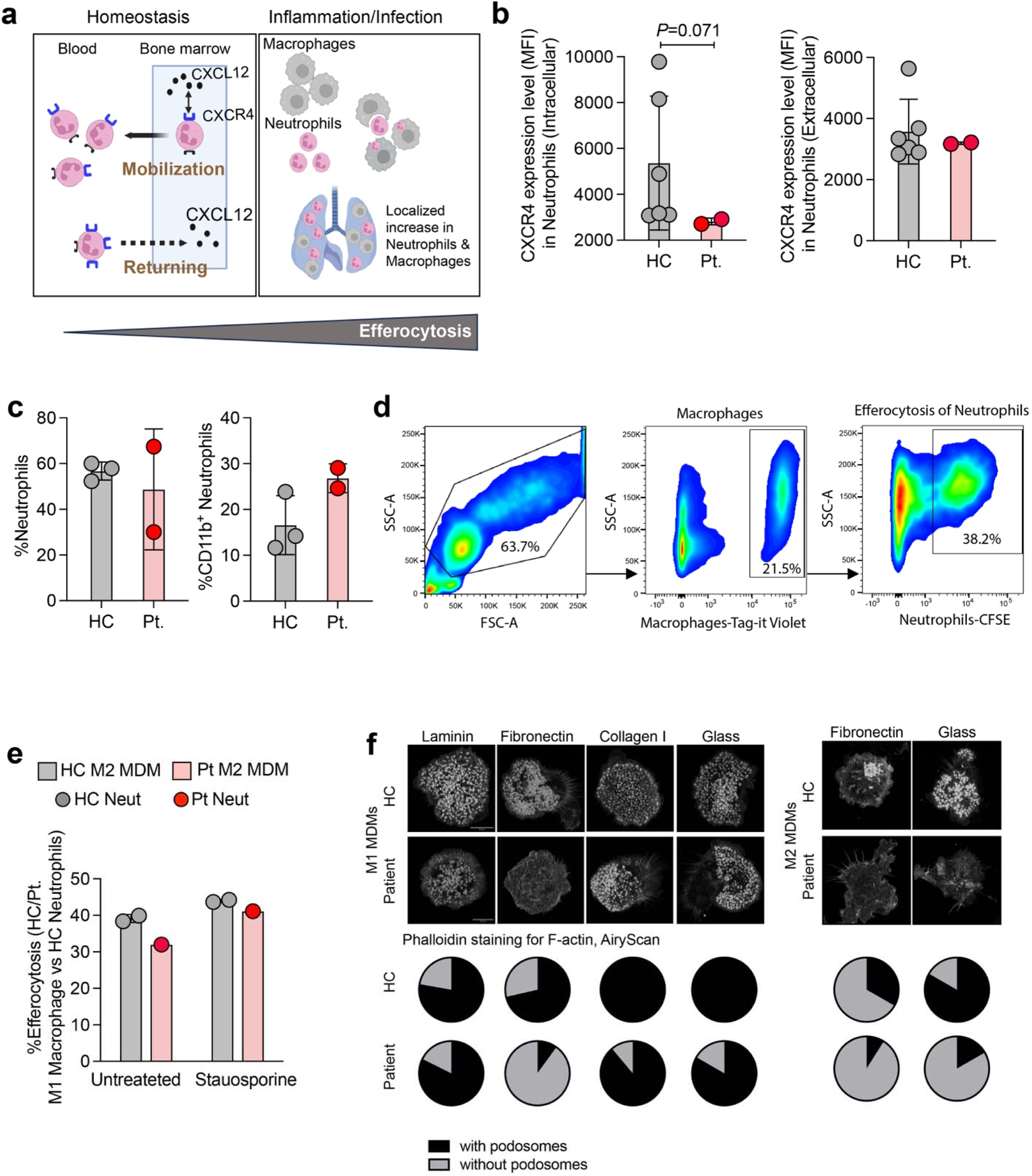
Impaired neutrophil clearance and macrophage membrane dynamics in dysferlin-mutant cells. **a,** Schematic illustrating neutrophil clearance during homeostasis and inflammation. During homeostasis, aged neutrophils are removed from the circulation in the bone marrow and are characterized by increased surface CXCR4 expression. During infection or inflammation, exemplified here in the lung, neutrophils are cleared by macrophage-mediated efferocytosis. **b,** Total cellular CXCR4 expression was reduced in patient neutrophils compared with healthy controls, whereas surface CXCR4 expression remained unchanged. **c,** Neutrophil frequencies in whole blood from patients and healthy controls. **d,** Gating strategy used for the co-culture efferocytosis assay. **e,** Adherent PBMC-derived monocytes were cultured with GM-CSF for 7 d to generate M1-like monocyte-derived macrophages. Efferocytosis assays demonstrated no significant difference between patient-derived and HC M1 macrophages following co-culture with neutrophils. **f,** M1 monocyte-derived macrophages were plated on glass or on chambers pre-coated with laminin, fibronectin or collagen I to assess podosome formation. F-actin was visualized by phalloidin staining and imaged using confocal microscopy with Airyscan

**Extended Data Fig 5.**
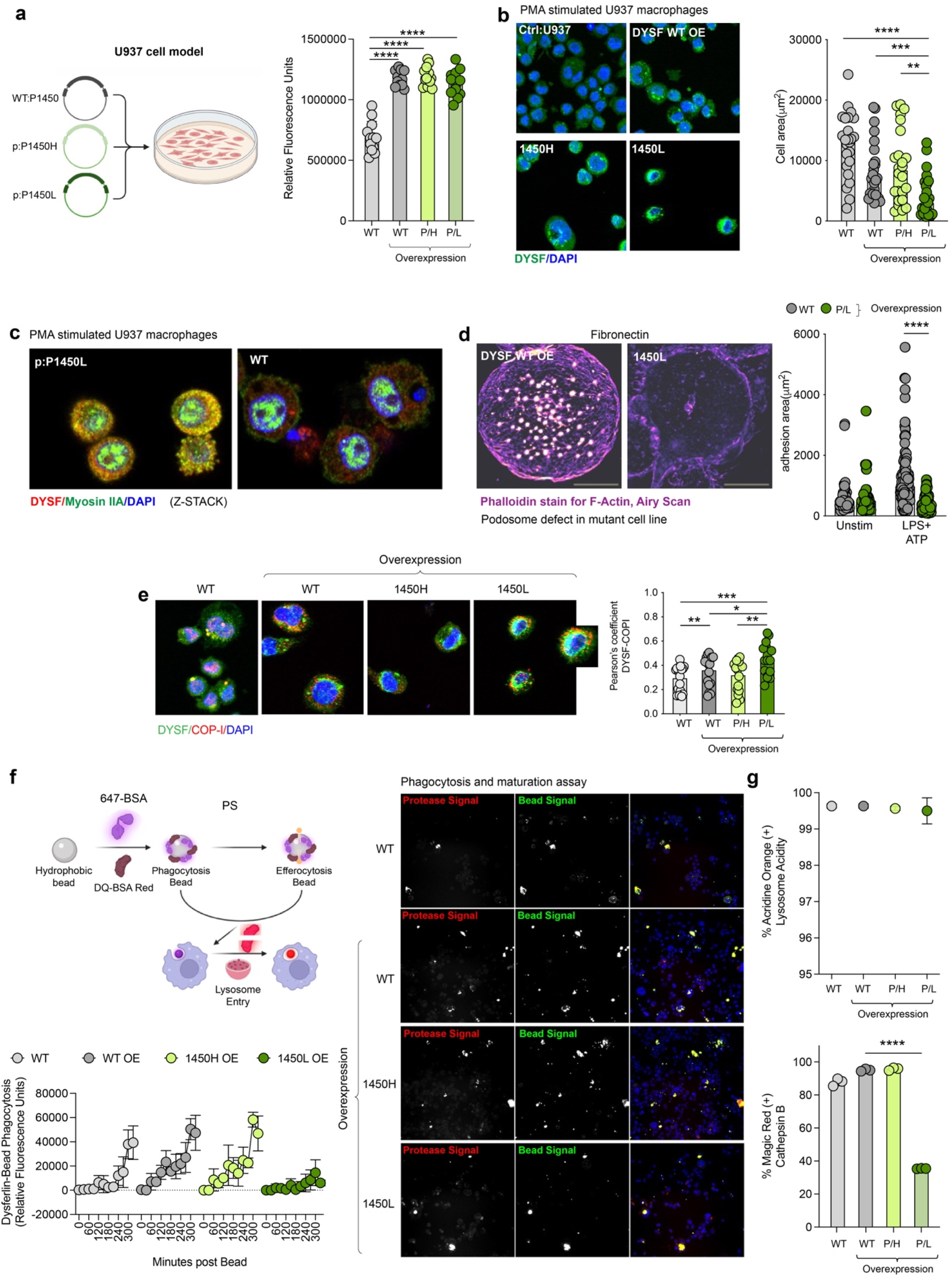
Dysferlin-mutant U937 cells recapitulate defects in cytoskeletal organization, phagocytosis and lysosomal function. **a,** Quantification of dysferlin fluorescence intensity in wild-type (WT), dysferlin-overexpressing and mutant dysferlin-expressing U937 cells. **b,** WT, dysferlin-overexpressing and mutant dysferlin-transfected U937 cells were differentiated with PMA (30 ng /ml) and M-CSF (100 ng ml/ml) for 2 days, rested for 24 h and stimulated with L+A. Cells were stained with anti-dysferlin antibody (green), and nuclei were counterstained with DAPI. **c,** Representative immunofluorescence images of WT and mutant dysferlin-transfected U937 cells co-stained for dysferlin and myosin. **d,** PMA-differentiated WT and mutant U937 cells were plated on glass or fibronectin-coated chamber slides to assess podosome formation. F-actin was visualized by phalloidin staining and imaged using Airyscan microscopy. **e,** U937 cells were differentiated with PMA (30 ng /ml) and M-CSF (100 ng /ml) for 2 days, rested for 24 h, stimulated with L+A and stained with anti-dysferlin antibody (green) and anti-COPI antibody. Nuclei were counterstained with DAPI. **f,** The U937 transfection model recapitulates the phagocytic defects observed in dysferlin-mutant cells. In vitro assessment of phosphatidylserine (PS)-mediated efferocytosis and phagosome maturation demonstrated that the p.P1449L dysferlin variant failed to enhance efferocytosis compared with dysferlin-overexpressing cells and exhibited a marked defect in phagosome maturation. **g**, Lysosomal activity was assessed by acridine orange staining. Cathepsin B activity was quantified using the Magic Red assay.

**Extended Data Fig 6.**
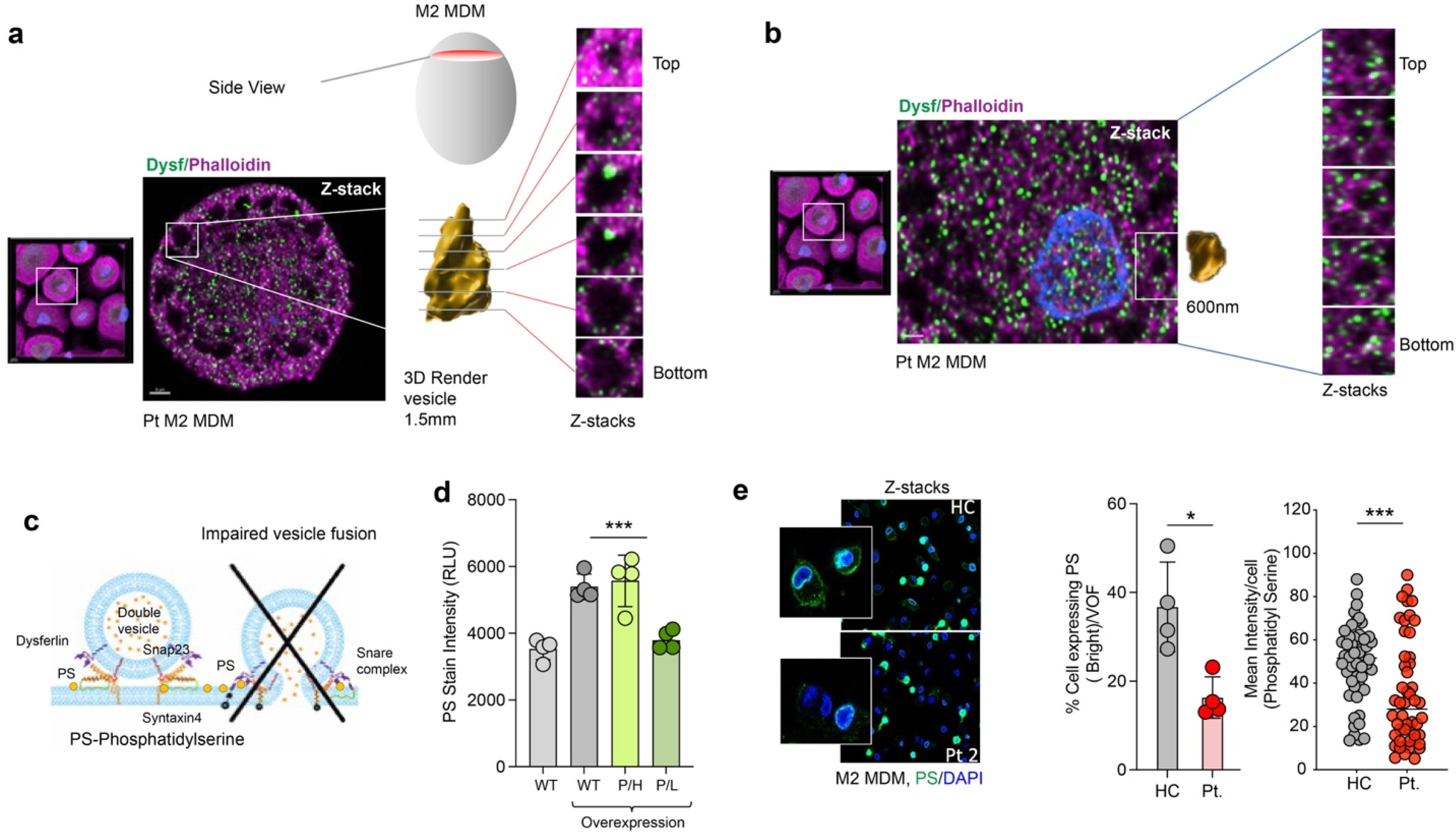
Dysferlin mutation alters vesicle architecture and phosphatidylserine distribution. **a,b,** Monocytes were cultured for 7 d and differentiated into M2-like macrophages. Cells were stained for dysferlin (DYSF), F-actin (phalloidin) and nuclei (DAPI). Representative immunofluorescence images show DYSF (green), phalloidin (purple) and DAPI (blue). Vesicular structures were segmented and reconstructed in three dimensions using Imaris v10.2.0. **c,** Schematic presentation of vesicle fusion. **d,** WT, dysferlin-overexpressing and mutant dysferlin transfected U937 cells were differentiated with PMA (30 ng /ml) and M-CSF (100 ng ml/ml) for 2 days, rested for 24 h and stimulated with LPS and ATP (L+A). Cells were stained with anti-phosphatidylserine antibody. **e,** Monocytes from patients and healthy controls were cultured for 7 d and differentiated into M2-like monocyte-derived macrophages. Cells were stained with an anti-phosphatidylserine antibody, and phosphatidylserine fluorescence intensity was quantified using Imaris software.

**Extended Data Fig 7.**
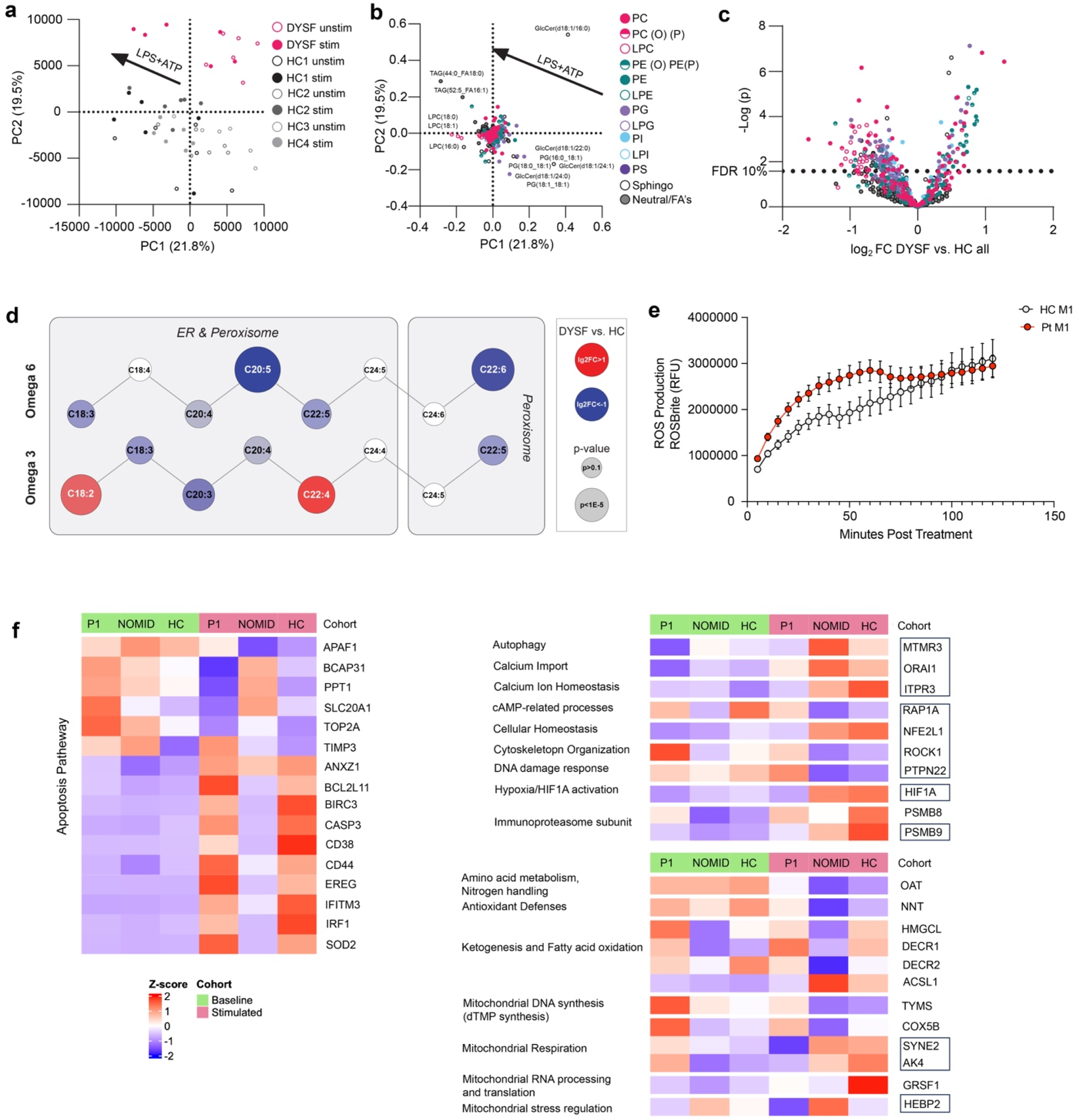
Lipidomic and transcriptomic profiling reveals altered lipid metabolism and cellular stress responses in dysferlin-mutant cells. **a,** Principal component analysis of Pareto scaled lipidomic data from PBMCs. A shared vector of variance associated with LPS + ATP stimulation is indicated. **b,** Corresponding loading plot for the PCA shown in **a**. Lipids are colored according to lipid class, and selected lipids of interest are labeled. **c,** Full volcano plot showing the significance and magnitude of lipid changes between DYSF patient PBMCs and healthy volunteers under unstimulated conditions. Plot corresponds to main Figure 4a-c. A-line indicating the Benjamini-Hochberg FDR < 10% cut-off is shown. **d,** Pathway map showing elongation and iterative desaturation of omega-3 and omega-6 polyunsaturated fatty acids. The average log2 fold-change of lipids containing each acyl chain in DYSF patient PBMCs compared with healthy controls is represented by node color. The average log10 *P* value of lipids in each group is represented by node size. Subcellular localization of each process is indicated by boxed regions. **e,** Total reactive oxygen species levels were measured in patient-derived M1 monocyte derived macrophages compared with healthy controls following LPS + ATP stimulation. **f,** Bulk RNA-sequencing analysis of apoptosis, mitochondrial and interferon-response pathway genes in patient P1 (*n* = 1), NOMID controls (*n* = 3) and healthy controls (*n* = 2) at baseline and following LPS and ATP stimulation of M2 macrophages demonstrate impaired adaptive responses involving interconnected stress-response networks. DYSF-mutant M2 MDMs failed to induce genes involved in autophagy and calcium homeostasis (*MTMR3, ORAI1, ITPR3*), as well as key regulators of oxidative and metabolic stress adaptation (*NFE2L1* and *HIF1A*). Impaired stress adaptation was further accompanied by dysregulation of mitochondrial homeostatic pathways, including antioxidant defense (*NNT*), fatty acid oxidation and ketogenesis (*HMGCL, DECR1, DECR2, ACSL1*), mitochondrial DNA synthesis (*TYMS*), respiratory chain function (*COX5B, AK4, SYNE2*), mitochondrial RNA processing and translation (*GRSF1*), and mitochondrial stress regulation (*HEBP2*). Differential pathway activity is shown as log2 fold change relative to healthy controls.

**Extended Data Fig 8.**
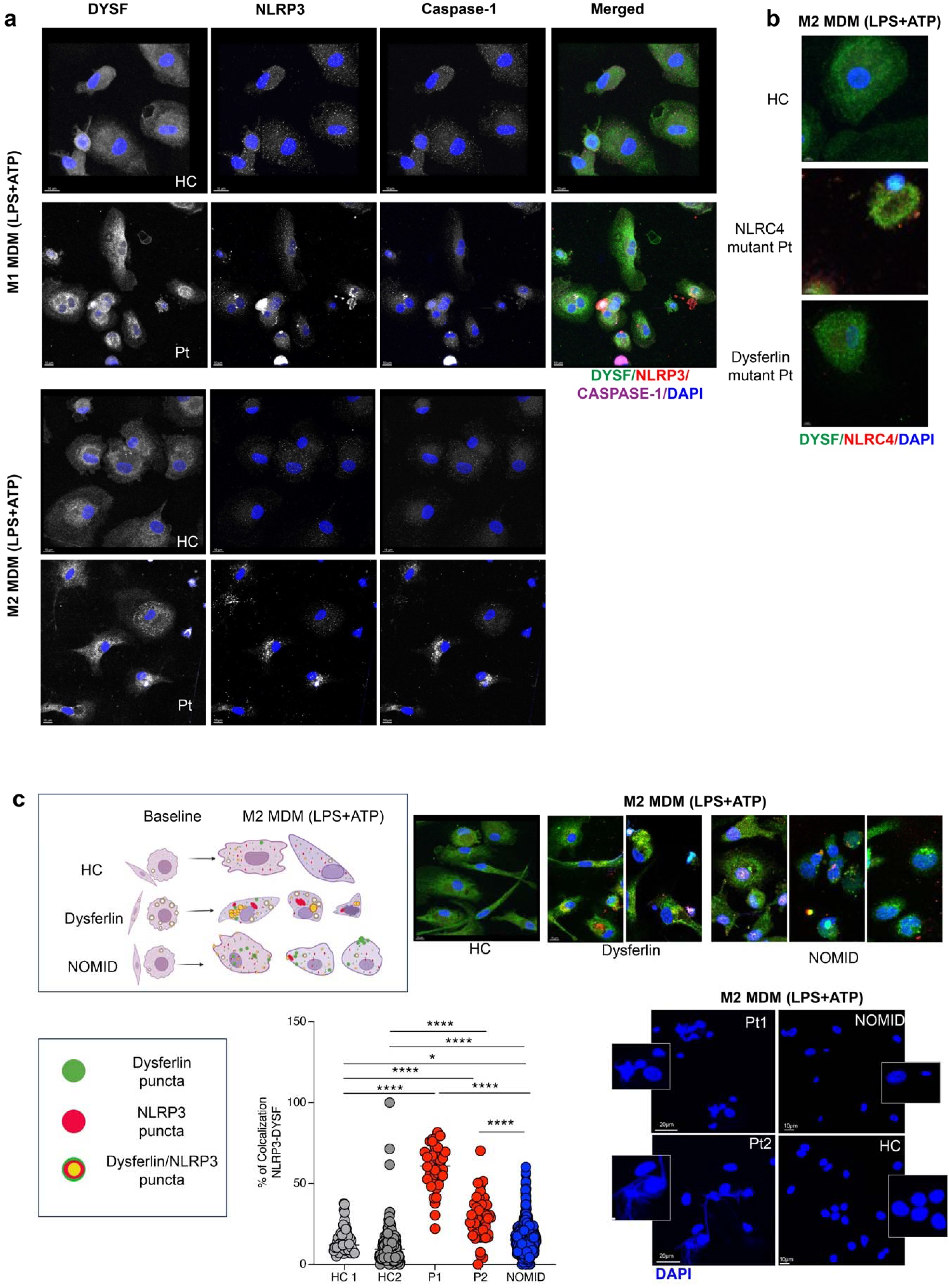
Dysferlin colocalizes with NLRP3 and caspase-1 but not NLRC4 in patient-derived macrophages. **a,** Monocytes were cultured for 7 d and differentiated into M1/M2-like monocyte-derived macrophages (MDMs). Cells were stained with antibodies against dysferlin (DYSF), NLRP3 and caspase-1, together with DAPI. Representative immunofluorescence shows dysferlin (DYSF, green), NLRP3 (red) and caspase-1 (purple) expression and localization in patient-derived M1 MDMs with individual channels in grey, also individual staining channels shown in grey for DYSF (green), NLRP3 (red), caspase-1 (purple) and nuclei (DAPI) for M2 MDM. **b,** M2-like MDMs were stained with antibodies against NLRC4 (red) and dysferlin (green). **c,** M2 MDMs were stained with antibodies against dysferlin (DYSF) and NLRP3, together with DAPI. Representative immunofluorescence images show distinct pattern of NLRP3 in stimulated M2 MDMs also revealing dysmorphic nuclei that were predominantly seen in dysferlin-mutant M2 MDMs. Percent colocoalization(NLRP3-DYSF) was measured for individual cells and is described in supplementary method section.

**Extended Data Fig 9.**
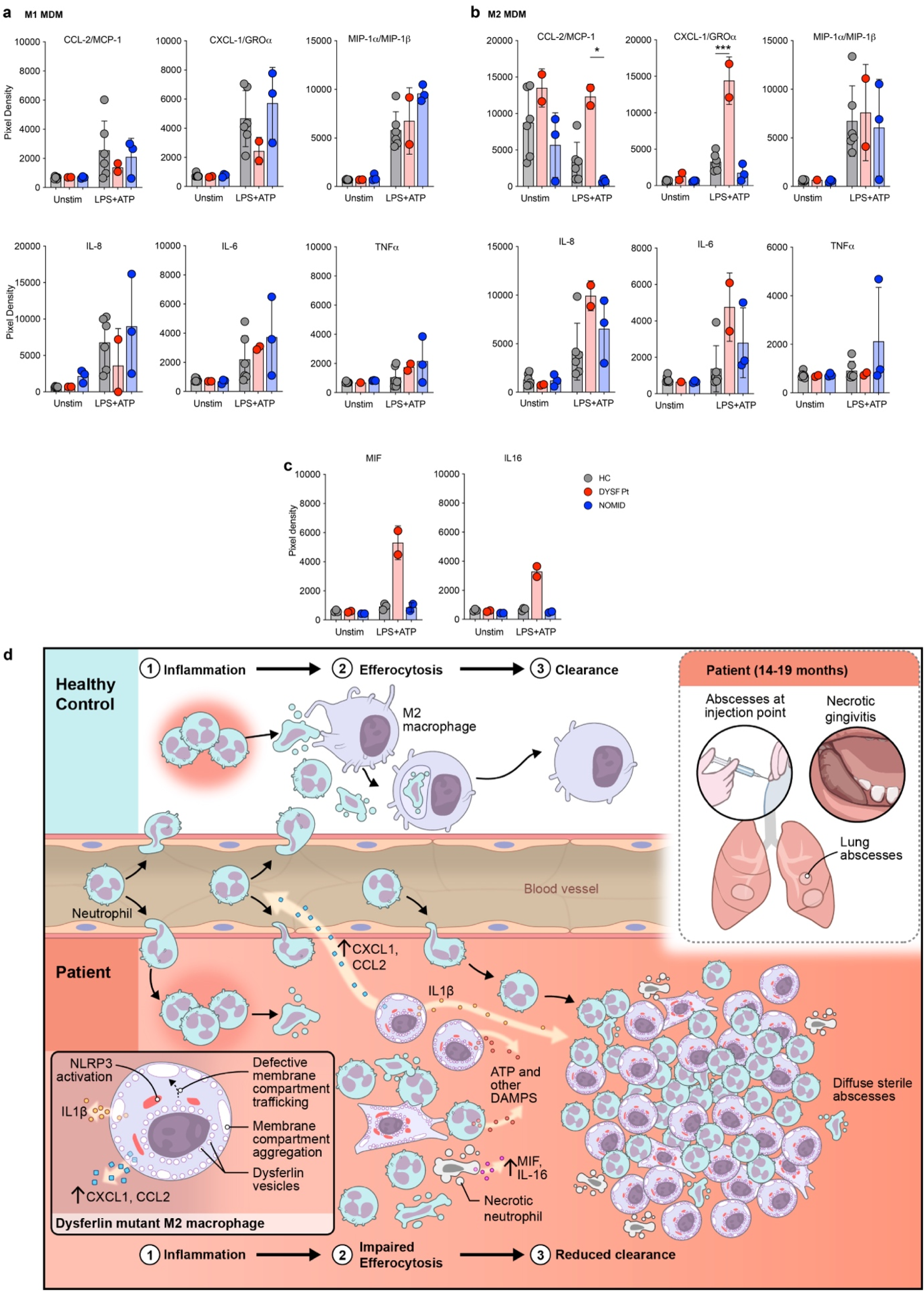
Dysferlin–NLRP3-positive extracellular puncta associated with IL-1-driven inflammatory signaling. **a,b,** Cytokine and chemokine production by M1-like macrophages (**a**) and M2-like macrophages (**b**) after stimulation with LPS, 1 μg/ml, and ATP, 3 mM, for 24 h. Relative cytokine and chemokine levels in culture supernatants were analyzed by antibody array, and spot density was quantified using ImageJ. Each symbol represents an individual subject: healthy controls (n = 6), patient (n = 2), patients with NOMID (n = 3). **c,** Neutrophils were isolated from patient and stimulated with LPS and ATP for 3 hours and supernatant were collected. Relative cytokine and chemokine levels in culture supernatants were analyzed by antibody array, and spot density was quantified using imaje J. (*n* = 3), patient (*n* = 2), patients with NOMID (*n* = 2) Bars show the mean ± SD **d,** Model of IL-1-associated danger signalling and NLRP3 activation in dysferlin-mutant macrophages. Defective efferocytosis promotes neutrophil accumulation, impaired inflammatory clearance and sterile abscess formation.

## Notes

### Competing Interest Statement

The authors have declared no competing interest.

### Author Declarations

The Institutional Review Board of the National Institutes of Health gave ethical approval for this work under the National Institute of Allergy and Infectious Diseases Natural History Study protocol (Number: 17-I-0016 (NCT02974595)).

