## Supplementary material for "Novel gain-of-function mutation in dysferlin causes vesicle trafficking defect and IL-1 mediated autoinflammation": 0.4 Final FINAL Supplementary information MedRxiv.docx

**CONTENTS**

**I. Supplementary Methods**

**II. Supplementary Tables**

**III. Supplementary Notes**

**IV. Supplementary Video**

**Supplementary References**

**Supplementary Notes (including notes clarifying statistical analyses, acknowledgements, grant or other numbers)**

I. **Supplementary Methods:**

***Study participants***

All research investigations were conducted under protocol 17-I-0016/NCT02974595, that was approved by the NIAID and NIAMS/NIDDK Institutional Review Boards. Patients were referred to the NIH between 2013 and 2015. Written informed consent was obtained from the parents of all patients/healthy controls involved, and assent was obtained from the patients/healthy controls were indicated. Both patients were referred for unexplained systemic inflammation and sterile lung abscesses and underwent trio analysis at the NIH Clinical Center. Clinical, laboratory and imaging data and unstained slides from clinically indicated skin, gastrointestinal and liver biopsies were obtained and stained for research markers. The data collection period for some parameters is from birth to April 2022 for both patients. The NIH protocol enrolls parents and healthy siblings as controls. The control experiments were conducted with blood from the patients' parents and or controls from the blood bank approved under the protocol. The authors affirm that the patients (or their parents/legal guardians) provided written informed consent for publication of the images in the Supplementary Fig. 1 and the medical information included in this paper. Anakinra was initially started as clinical treatment. Canakinumab was later initiated once patients were in inflammatory remission and had normalized their inflammatory parameters on an off-label basis. The study was reported according to CARE guidelines and conducted in compliance with the Declaration of Helsinki principles.

**Annotation of the disease-causing variant throughout the manuscript:**

**Nomenclature note:** The mutation is designated p.P1449L throughout this manuscript according to *DYSF* **isoform 7** *(DYSF-7)*, the predominant isoform expressed in myeloid cells and the isoform used for functional studies. The same amino acid substitution corresponds to p.P1432L in the canonical muscle isoform (**isoform 1,** *DYSF-1*) and p.P1450L in **isoform 13** (*DYSF-13*), that was used in the U937 tranfection studies. For consistency and biological relevance to the disease mechanism described here, the p.P1449L nomenclature is used throughout unless indicated otherwise.

***Generation of Dysferlin mutant U937 cell line***

Production of U937 cells stably expressing dysferlin mutants was performed by making MIG-series retroviral constructs. Briefly, virus was produced by transfecting 293T17 cells with a 3:1:0.1 ratio of EcoPak (Addgene), construct, and VSV-G (Addgene) using Mirus Lenti (). After 72 h, virus was collected and 0.45 um filtered and was directly applied to log-phase U937 cells. After 72 hours, cells were sorted by GFP expression using a FACS Aria and the upper quartile of expressors were use for subsequent experiments.

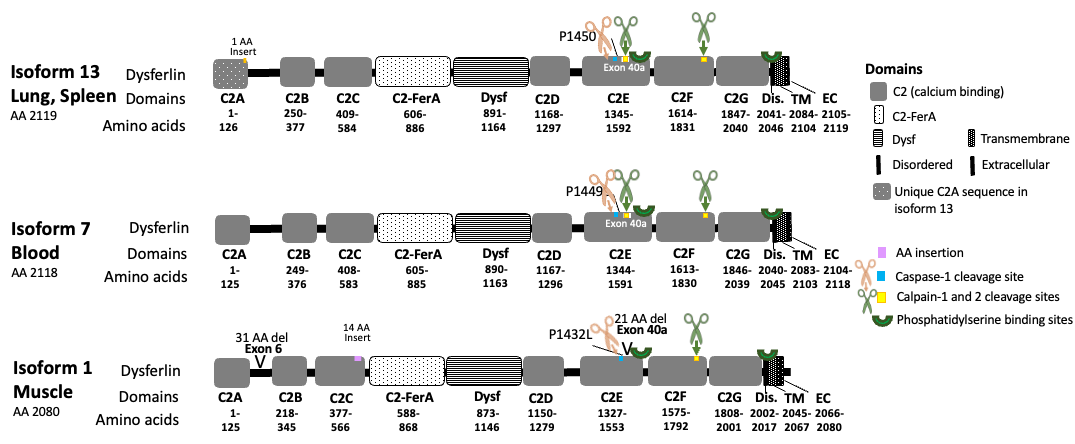

***Figure***

***Colocalization Analysis***

All analyses were restricted to voxels inside Imaris-segmented cell objects (Cells0 mask) background voxels were excluded. Two colocalization approaches were applied:

Cell-level Pearson/Spearman r  computed across per-cell mean intensities; quantifies whether high-green cells are also high-magenta. Fisher z-test compared between sample groups.Mann-Whitney U and Welch t-test (FDR-corrected) were applied across all metrics.

Costes automatic threshold colocalization([1]) **was done** for pixel-level analysis within cell voxels only; automatic threshold eliminates arbitrary intensity cutoffs. Manders M1, M2, and Overlap Coefficient computed at Costes thresholds.

II. **Supplementary Tables:**

**Supplementary Table 1. Features of the *DYSF* Mutation Detected in Patients 1 and 2.**

| Gene | *DYSF* |
| --- | --- |
| pLI | 0.00 |
| Transcript | ENST00000409582.7/ NM_001130981.2 |
| Chr coordinates (GRCh38) | Chr2:71612768 C>T |
| HGVSc | c.4346C>T (*DYSF-1*)* |
| HGVSp | p.P1449L (*DYSF-7*)* |
| Exon | 39/56 |
| Inheritance mode | De novo dominant |
| rsID | NA |
| gnomAD AF | 0 |
| AllofUs AF | 0 |
| GERP score | 3.72 |
| CADD PHRED | 22.9 |
| REVEL | 0.33 |
| AlphaMissense | 0.196 |
| MetaSVM_prediction | Tolerated |
| MutationTaster_prediction | Disease causing |
| PROVEAN_prediction | Damaging |
| Polyphen2_HDIV_prediction | Benign |
| SIFT_prediction | Tolerated |
| FATHMM | Tolerated |
| BLOSUM62 | -3 |
| MSC-CADD damage prediction | High impact |
| EVEclass90 prediction | Benign |
| Present in COSMIC | No |

*Dysferlin isoform used to annotate AA change.

- pLI: probability of loss-of-function intolerance. *LYN* is intolerant to haploinsufficiency as indicated by a “probability of being loss-of-function intolerant” [pLI] score of 1.0, on a range of 0.0 to 1.0, with a higher score indicating a greater degree of intolerance for loss-of-function variants in healthy persons.
- HGVS: Human Genome Variation Society.
- gnomAD_AF: Genome Aggregation Database (gnomAD) v.2.1.1 minor allele frequency for all populations
- GERP: Genomic Evolutionary Rate Profiling. Score threshold of 2 or greater indicates truly constrained sites.
- CADD_phred: CADD Whole-genome Combined Annotation-Dependent Depletion (CADD) scores are based on conservation metrics, functional genomic data, transcript information, and protein-level scores (e.g. SIFT, and PolyPhen). Higher CADD_raw and CADD_phred scores indicate that a variant is more likely to be deleterious, CADD_phred scores >=10 predict that a variant is amongst the 10% most deleterious of all possible substitutions, CADD_phred scores >=20 predict that a variant is amongst the 1% most deleterious of all possible substitutions.[2]
- REVEL (Rare Exome Variant Ensemble Learner) is an ensemble method for predicting the pathogenicity of missense variants based on a combination. [3]
- AlphaMissense, is an adaptation of AlphaFold fine-tuned on human and primate variant population frequency databases to predict missense variant pathogenicity. [4].
- MetaSVM: Radial kernel support vector machine network based on nine prediction scores and allele frequencies in 1000G.[5]
- PROVEAN (Protein Variation Effect Analyzer) is a software tool that predicts whether an amino acid substitution or indel has an impact on the biological function of a protein.[6]
- PolyPhen2_HDIV: Polymorphism Phenotyping v2.[7]
- SIFT: Sorting Intolerant From Tolerant.[8]
- BLOSUM62: BLOcks of Amino Acid SUbstitution Matrix, a positive BLOSUM62 score means that the amino acid substitution is conservative, the two amino acids are biochemically similar
- MSC-CADD: Mutation Significance Cutoff-Combined Annotation Dependent Depletion
- COSMIC: Catalogue of Somatic Mutations in Cancer

III. **Supplementary Notes:**

**Supplementary Note 1**

**Case descriptions**

Additional clinical information regarding the cases is available from the corresponding author upon reasonable request.

**Supplementary Note 2**

**Modeling of Dysferlin (Isoform 7):**

We generated five structural models of dysferlin isoform 7 (UNIPROT ID O75923-7) from its sequence using the AlphaFold software (Jumper et al., 2022) installed on the Biowulf cluster at the National Institutes of Health. We took the top-scored conformation along with estimates of prediction reliability (pLDDT) and predicted aligned error (PAE), as described elsewhere (Evans et al., 2021). Briefly, pLDDT indicates the quality of prediction for individual residues in a protein domain, while PAE is governed by the prediction of the interaction between domains, consequently indicating the reliability of the relative position of domains in a multidomain protein.

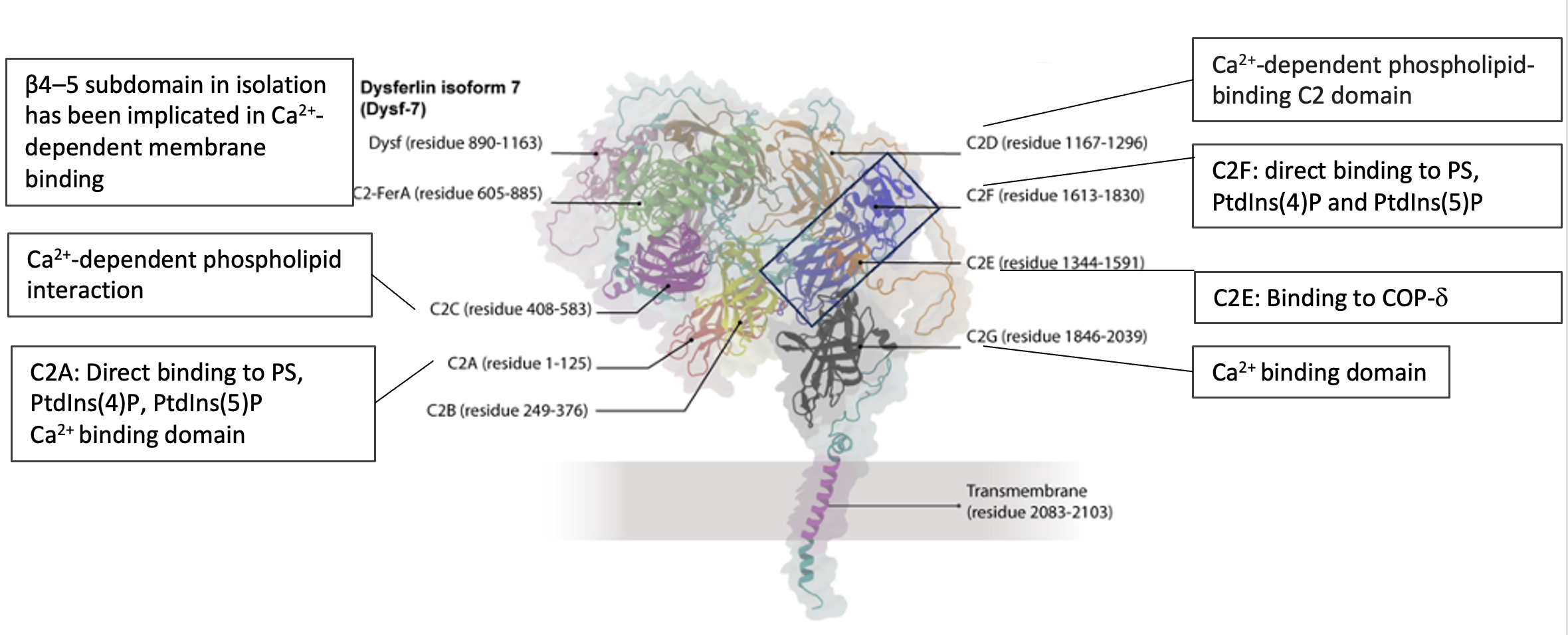

***Model of dysferlin isoform 7 (Dysf-7) generated by AlphaFold***. Different domains of Dysf-7 are shown in different colors. The definition of the dysferlin domains is taken from the literature (Dominguez, 2022), where the domain architectures of the ferlin protein family have been redefined according to the structural motifs of the proteins. Dysf-7 domains, determined according to the above domain definitions of ferlin protein family, are shown in different colors - C2A (residue 1-125) in red, C2B (residue 249-376) in yellow, C2C (residue 408-583) in purple, C2-FerA (residue 605-885) in lime, Dysf (residue 890-1163) in Mauve, C2D (residue 1167-1296) in ochre, C2E (residue 1344-1591) in orange, C2F (residue 1613-1830) in blue, C2G (residue 1846-2039) in black, and transmembrane (residue 2083-2103) in magenta. The remaining residues are shown in cyan. The proline in the mutation position, residue 1449, is shown in orange in the ball and stick model. Two other proline residues, P1591 and P1601, near P1449, are shown in orange in the ball and stick model. These three proline residues have the potential to form a proline-proline stack. Proline-rich regions, through stacking, usually facilitate bringing proteins together so that subsequent interactions required for protein function become probable

Dysferlin has a single transmembrane domain at the C terminus and a long N-terminal cytoplasmic region that contains six C2 domains. C2 domains are a common feature of the synaptotagmin family of proteins regulating vesicular traffic and membrane fusion events through calcium-dependent interactions with phospholipids and proteins [9, 10]

C2A domain: can directly bind to PS, PtdIns(4)P, PtdIns(5)P; and has a Ca^2+^ binding domain

C2C domain: Ca^2+^-dependent phospholipid interaction

C2FerA and Dysf domains: The β4–5 subdomain in isolation has been implicated in Ca2+-dependent membrane binding

C2D domain: Ca^2+^-dependent phospholipid-binding C2 domain

C2E domain: Binding to COP-d

C2F domain: direct binding to PS, PtdIns(4)P and PtdIns(5)P

C2G domain: Ca^2+^ binding domain

Transmembrane region

*Direct interaction with*: Snapin-23, Vamp-3, Syntaxin-4 [11]

**Supplementary Note 3**

RNA-seq expression levels, shown as RPKM, are plotted for **DYSF** (dysferlin), **LAMP1** (CD107a), and genes encoding COPI coatomer subunits across unstimulated and stimulated M1 and M2 macrophages in patient compared to HC and NOMID.COPI is composed of seven core coatomer subunits: α-COP, β′-COP, ε-COP, β-COP, δ-COP, γ-COP, and ζ-COP. These subunits assemble in the cytoplasm into a stable heptameric coatomer complex that is recruited to Golgi membranes to mediate COPI-coated vesicle formation and retrograde trafficking within the Golgi and from the Golgi to the endoplasmic reticulum. DYSF mutant(n=1) M1 or M2-like MDMs are in red, NOMID(n=3) MDMs are in blue and controls(n=2) MDMs are in in grey.

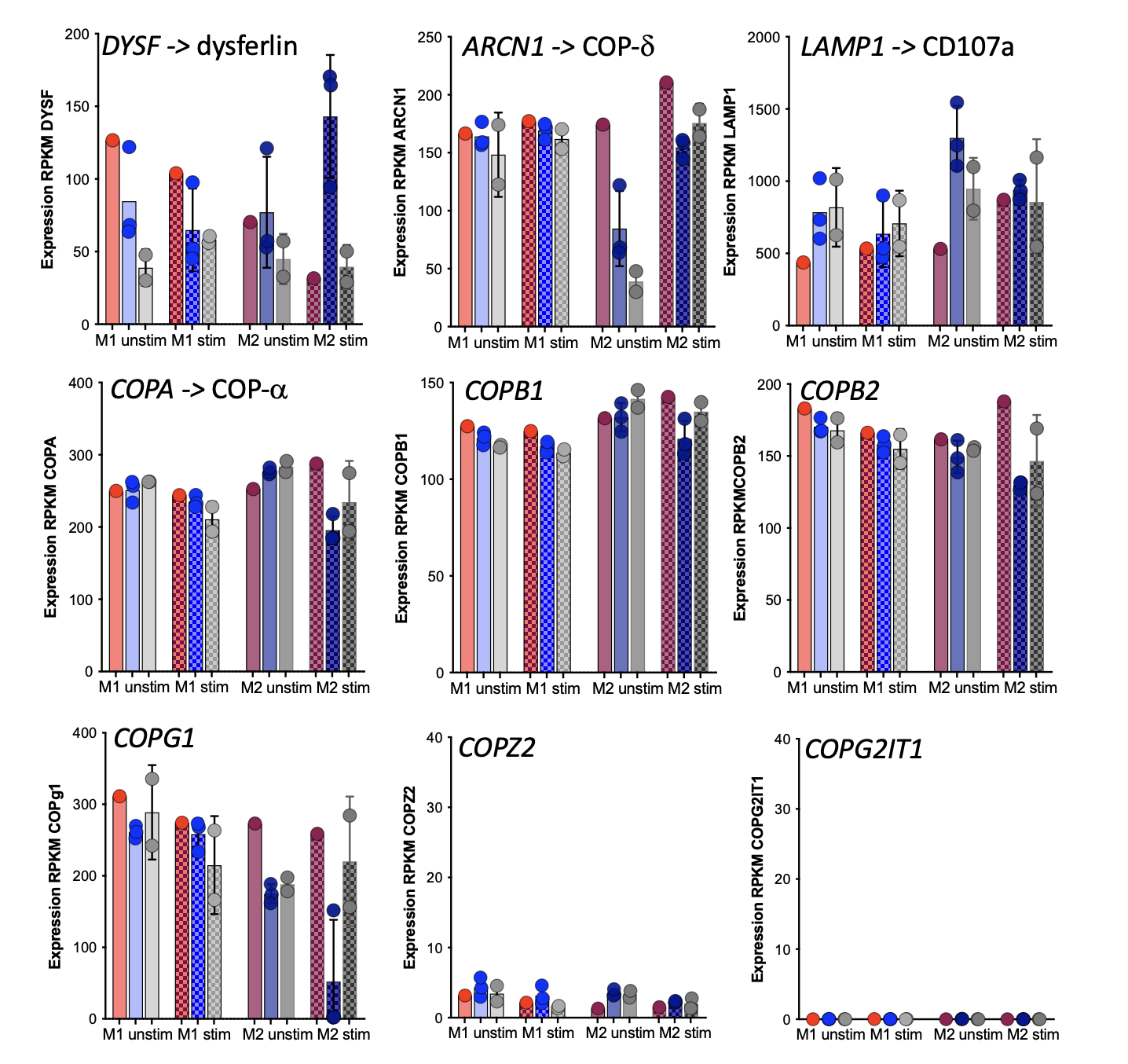

Figure**:****Relative mRNA expression in M2 MDMs (monocyte derived macrophages).** Bar graph showing transcriptional expression levels of selected genes/markers in M2 MDMs across experimental groups. Bars represent mean expression values, and individual points indicate biological replicates. Error bars indicate variability among replicates.

**Supplementary Note 4**

We observed dysferlin association with enlarged vesicle-like structures resolvable by light microscopy, as well as perinuclear dysferlin-positive compartments, although the smaller perinuclear structures cannot be definitively resolved at the ultrastructural level**.** (Extended Data Fig 6a,b)
