## Supplementary material for "Novel gain-of-function mutation in dysferlin causes vesicle trafficking defect and IL-1 mediated autoinflammation": Supplementary videos.pptx

### Slide 1
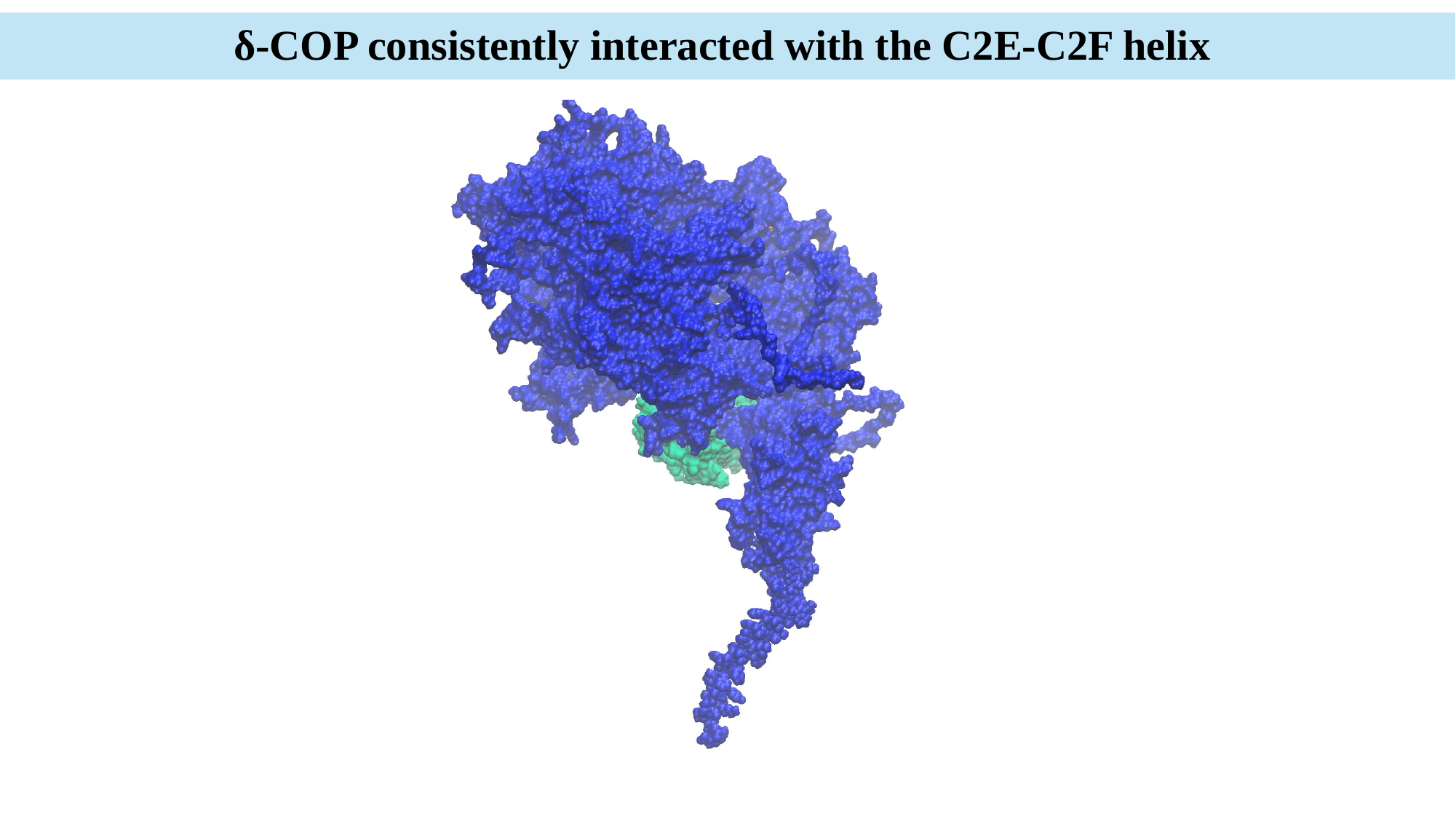

δ-COP consistently interacted with the C2E-C2F helix

### Slide 2
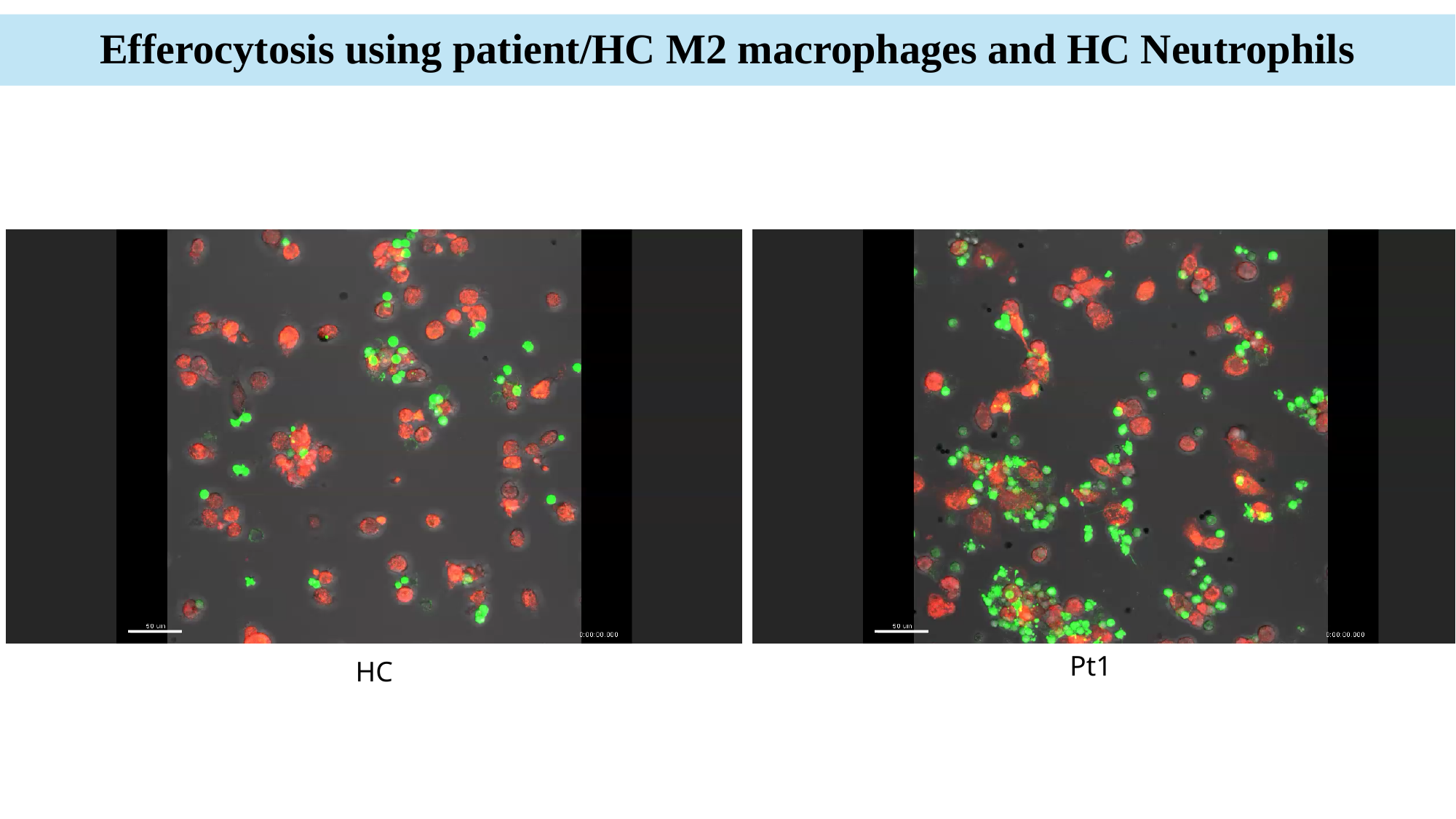

Efferocytosis using patient/HC M2 macrophages and HC Neutrophils
#### Efferocytosis using patient/HC macrophages and HC Neutrophils
Pt1
HC
